# Predicting Four-Class Residual Cancer Burden (RCB) Before Neoadjuvant Chemotherapy in Breast Cancer: Block-Structured Model Comparison, Selection Criterion and Decision Threshold

**DOI:** 10.64898/2026.08.16.26360557

**Authors:** Yusuf Kagan Dagdeviren, Huseyin Salih Semiz, Erdinc Hakan Inan, Heves Yaren Karakas, Merih Guray Durak, Nazimcan Tezel, Mert Can Sevindik, Petek Ballar Kirmizibayrak, Recep Bekis

**Affiliations:** Hospital Pharmacy, Dokuz Eylul University Hospital, Inciralti Mah. Mithatpasa Cad. No: 1606, 35330 Balcova, Izmir, Turkiye; Institute of Oncology, Department of Preventive Oncology, Dokuz Eylul University, Izmir, Turkiye; Department of Radiology, Antalya Training and Research Hospital, Antalya, Turkiye; Department of Pathology, Dokuz Eylul University, Izmir, Turkiye; Department of Biochemistry, Faculty of Pharmacy, Ege University, Izmir, Turkiye; Department of Nuclear Medicine, Dokuz Eylul University, Izmir, Turkiye

**Keywords:** Breast neoplasms, Neoadjuvant therapy, Residual cancer burden, Machine learning, Model selection, Explainable artificial intelligence

## Abstract

**Background and Objective:** Residual cancer burden (RCB) after neoadjuvant chemotherapy (NAC) stratifies prognosis more finely than binary pathologic complete response, yet prediction models routinely collapse this four-tier scale. We predicted all four classes from pre-treatment data and examined how the selection criterion and decision threshold determine the outcome in an imbalanced multiclass problem.

**Methods:** In a single-centre retrospective cohort of 328 patients treated with NAC followed by surgery, 64 candidate variables were organised into six thematic blocks; four were excluded, two for missingness and two because they are not known before NAC, leaving 60 features. A 17-variable radiology model (Model R) was compared with a 60-variable multimodal model (Model ALL) across 100 stratified splits. Within each split, selection was repeated by five-fold inner cross-validation on the training data using the mean of macro AUC and macro-F1; the held-out test set was used once, after selection. Reporting follows TRIPOD+AI.

**Results:** RCB classes were distributed as RCB-0 27.4% (n=90), RCB-I 10.4% (n=34), RCB-II 43.6% (n=143) and RCB-III 18.6% (n=61). Across 100 splits the median test AUC was 0.823 (IQR 0.803–0.838) for Model ALL and 0.773 (0.746–0.803) for Model R, Model ALL being superior in 88% of splits (*d_z_*=1.18), in 84% for macro-F1 and in 98% for ordinal agreement (median quadratic weighted kappa 0.504 versus 0.260; *d_z_*=1.69). For the most frequently selected configuration, class-wise precision ranged from 0.300 to 1.000, sensitivity from 0.250 to 0.722 and specificity from 0.703 to 1.000. On one fixed split Model R fell at the 98th percentile of its own distribution and Model ALL at the 31st, reversing the ranking. The default 0.50 threshold drove sensitivity to zero at the intensification boundary (RCB-III versus the rest) in 48 of 100 splits for Model ALL and 51 for Model R; a threshold selected on training data raised median sensitivity to 0.833 and lowered specificity to 0.537.

**Conclusions:** The multimodal model is consistently superior in discrimination and markedly so in ordinal agreement. The selection criterion and the decision threshold are separate design decisions, and evaluation resting on a single split can reverse the ranking of models.

## 1. Introduction

Neoadjuvant chemotherapy (NAC) offers a rare in vivo readout of how a breast tumour responds to systemic treatment, yet predicting the depth of that response before treatment begins remains unsolved. Guidelines emphasise biomarker-based risk stratification while acknowledging that single markers cannot capture the clinic’s layered reality [1–3].

NAC response is most often read through pathologic complete response (pCR), a binary endpoint that groups minimal and extensive residual disease under one label and that a large pooled analysis could not validate as a surrogate for survival [4,5]. Residual cancer burden (RCB) instead grades remaining invasive disease on a continuous index [6], and its association with long-term outcome is established across subtypes [7] and in 5161 pooled patients [8]. Machine-learning models can bring tumour biology, patient characteristics and imaging phenotype into one framework [9,10], and imaging-based models already perform strongly [11]; the question is what happens to model selection once a strong imaging signal is combined with biology and disease extent.

Although RCB is a four-tier scale, the prediction-model literature routinely narrows it: deep-learning approaches on magnetic resonance imaging dichotomise it into RCB-0/I versus RCB-II/III [12], models working from biopsy images predict only the presence of residual disease [13], and the most comprehensive system to date, in 1048 patients across four institutions, produces three groups and requires mid-treatment imaging [14]. Such narrowing hides the two boundaries at which adjuvant intensity is decided. To our knowledge, no model predicting all four classes separately from pre-treatment data alone has been reported.

Opening the outcome to four classes carries a methodological cost: events per class fall, single-split estimates become more sensitive to sampling variability, and model selection becomes fragile. A systematic review of 152 machine-learning prediction models found the analysis domain most often at high risk of bias [15]. Cross-validation is biased at small sample sizes [16], model selection is itself a source of bias [17], and when split-induced variability is ignored the comparison becomes one between splits rather than models [18].

The criterion on which selection rests is a separate question: discrimination alone says nothing about how errors distribute across subclasses, so in an imbalanced multiclass outcome a rule looking only at discrimination may prefer a model that ignores a sparse class, and by the same logic the default arg-max rule may be unsuitable there. Explainability is also a condition for clinical adoption [19,20].

We addressed four questions together: outcome resolution, predicting all four RCB classes from pre-treatment data; model selection, comparing the strongest single block with the integrated structure under selection confined to the training data; evaluation design, building a primary analysis on repeated rather than single splits; and the decision rule, testing the default threshold at clinically critical boundaries. Analyses follow TRIPOD+AI [21].

## 2. Methods

### 2.1. Study design and ethical framework

This was a single-centre, retrospective, observational cohort study of breast cancer patients who received NAC followed by surgery at Dokuz Eylul University Hospital, Izmir, Turkiye, between 2015 and 2024, drawing on the hospital information system, pathology records, the radiology archive and physical files. It was approved by the institutional Non-Interventional Clinical Research Ethics Committee (approval no. 2024/25-08; 17 July 2024); given the retrospective design, consent was waived and data were analysed after anonymisation.

### 2.2. Study population and outcome

Of 476 records screened, 328 entered the analytic cohort (Fig. 1); the largest exclusions were absence of NAC (n=46), unconfirmed invasive disease (n=33), no surgery after NAC (n=19) and baseline imaging or staging insufficient for feature extraction (n=16); disease stage was not an exclusion criterion. All 328 had confirmed invasive breast cancer and postoperative pathology allowing RCB to be calculated with the MD Anderson online calculator [6]: RCB-0 90 (27.4%), RCB-I 34 (10.4%), RCB-II 143 (43.6%) and RCB-III 61 (18.6%). The cohort is exclusively female; biological sex was defined as sex assigned at birth.

**Fig. 1.**
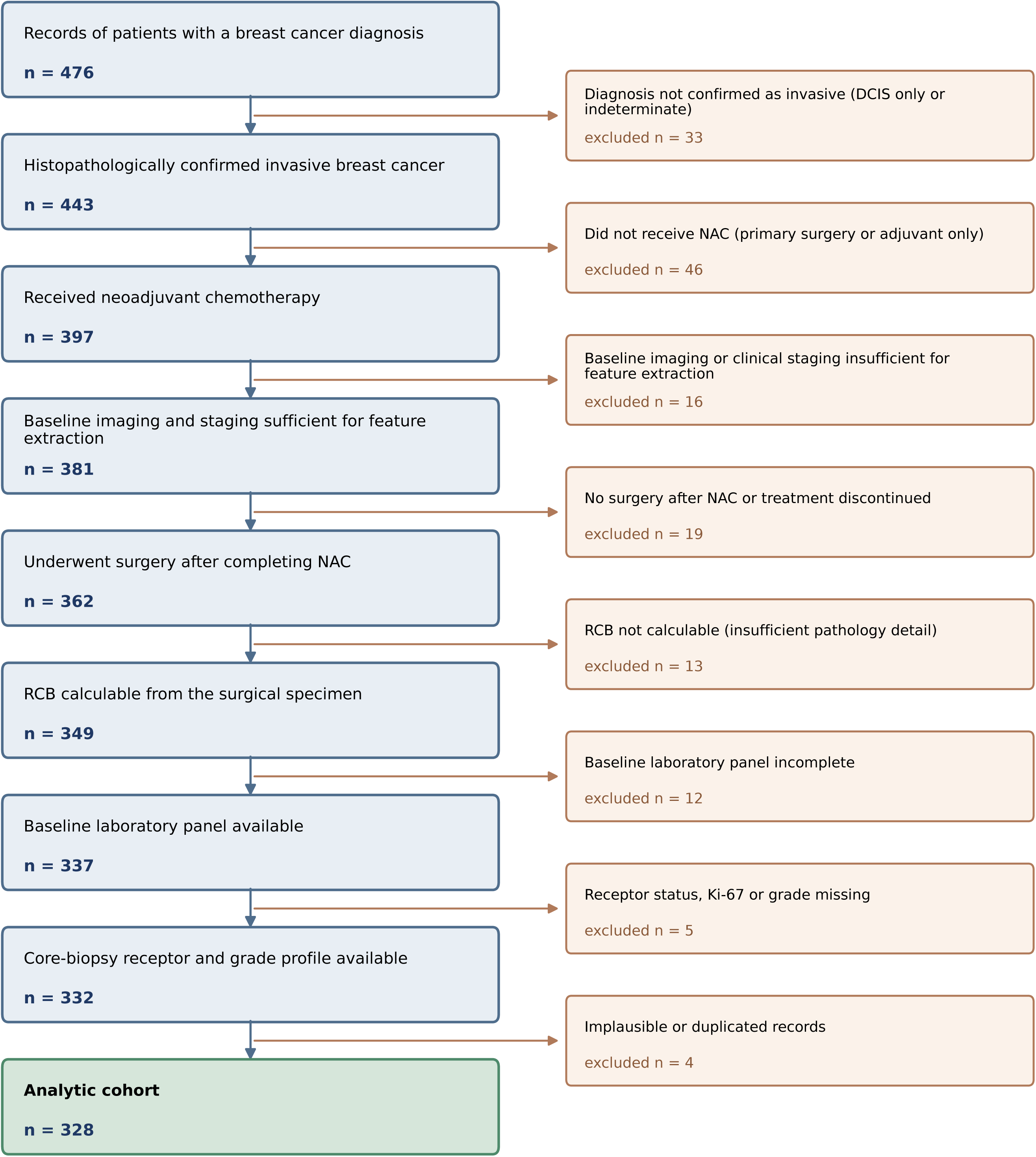
Flow of participants through the study. Of 476 records screened, 328 entered the analytic cohort.

All predictors were derived from data obtainable before the patient starts NAC: baseline imaging, pre-treatment core-biopsy pathology and receptor status, clinical stage, metastasis status and site, demographics, comorbidities and baseline laboratory values. Two treatment-process variables were deliberately excluded. Cycle intensity is known only once NAC is finished and carries reverse causality: a patient who fails to respond or develops toxicity stops treatment early, so incomplete treatment appears both as a consequence and as a predictor of poor response. Treatment regimen falls into a single category in 94.2% of the cohort. Excluding both did not lower performance (Section 3.6). The outcome came solely from the definitive surgical specimen.

Radiologic variables were abstracted from structured reports issued by the institutional radiology service and all reports were evaluated a second time, the second values used in the analyses. Across the 3,559 cells with a value in both evaluations, agreement was 89.0% (85.7– 92.6% at variable level). Cells undetermined at first abstraction were completed at the second evaluation, so no imputation was applied to this block.

### 2.3. Variable blocks and compared models

Candidate variables were organised into six thematic blocks (Supplementary Table S9); beyond this clinically motivated structuring, no automated feature selection was applied. Eleven configurations were evaluated, the main comparison being Model R, the 17 radiologic variables, against Model ALL, all blocks combined into 60 features. Four variables take a single value across the cohort (ALT, AST, LDH, TSH), so the effective count is 56 for Model ALL and 16 for Model R.

### 2.4. Missing data and preprocessing

In the pathologic, oncologic, demographic and biochemical blocks, missing values are cells coded as unknown. Missingness was assumed at random (Little’s test) and completed with a chained approach in a predefined order, each variable filled with a Random Forest classifier (100 trees) using only columns already complete or completed earlier in the chain. Two variables were excluded, E-cadherin (65.6% missing) and menopausal status (62.8%), both exceeding the 50% threshold and showing low masking-test accuracy (12.5% and 43.8%). Imputation preceded the split; the effect of this ordering is quantified in Section 3.6.

### 2.5. Modelling strategy, model selection and evaluation design

RCB was treated as a four-class classification problem. Three tree-based algorithms were evaluated — Random Forest [22], XGBoost [23] and LightGBM [24] — each with and without SMOTE applied only within training folds [25]. Hyperparameters were fixed at pre-specified values with no search, so that differences can be attributed to feature-block composition; the values are given in Supplementary Table S10.

The evaluation design was built to be consistent with the study’s own argument: a study contending that single-split performance is unreliable cannot rest its primary analysis on one split. The primary analysis therefore spans 100 stratified 80/20 splits. Within each, the six candidates were evaluated by five-fold cross-validation on that split’s training set, the candidate with the highest mean of macro one-vs-rest AUC and macro-F1 was selected, and the held-out test set was used once, after selection, so that the reported distribution incorporates both split and selection uncertainty. Each split was given to both models, with differences assessed by the paired Wilcoxon test and Cohen’s *d_z_*. Because the splits are drawn from the same patients and their training sets overlap, the primary inference rests on effect sizes rather than p values.

Combining two metrics was deliberate: selecting on discrimination alone can prefer models that lose the highest-risk class entirely. The rule was compared with inner-CV AUC alone and inner-CV macro-F1 alone (Section 3.2).

### 2.6. Evaluation metrics and additional analyses

Primary metrics were the macro one-vs-rest ROC AUC and macro-F1; accuracy and class-wise precision, sensitivity, F1-score, AUC and specificity were also computed. Because RCB is ordinal, quadratic weighted kappa, linearly weighted kappa, adjacent-class accuracy and mean absolute error were reported as well, together with two clinically defined boundaries: de-escalation (RCB-0/I versus RCB-II/III) and intensification (RCB-III versus the rest).

Analyses requiring a single model — two-level resampling, bootstrap confidence intervals, TreeSHAP explainability [20,26], calibration, decision curves and ablation — used the configuration selected most often across the 100 splits. In the ablation, each block was removed in turn and the paired change in test AUC measured across 20 splits. Calibration was assessed with Platt scaling and isotonic regression, clinical utility with decision curve analysis [27–29]. Thresholds were selected by cross-validation on training data only.

### 2.7. Statistical software

Analyses used Python 3.13 with scikit-learn 1.6.1, XGBoost 3.4.1, LightGBM 4.6.0, imbalanced-learn 0.14.2, SHAP 0.52.0, pandas 2.2.3, NumPy 2.1.3, SciPy 1.16.3 and statsmodels 0.15.0. The trained model object, feature list and class order are archived with the analysis code in an open repository with a persistent identifier.

## 3. Results

### 3.1. Cohort profile and comparison of configurations

Baseline characteristics by RCB class are given in Table 1. RCB-II was the largest class, followed by RCB-0, RCB-III and RCB-I; given this imbalance the modelling plan prioritised macro metrics and ordinal agreement over accuracy. Clinical stage, molecular subtype, receptor status, Ki-67, histologic grade and nodal status differed across RCB classes (p<0.001), whereas age group and TILs did not.

**Table 1.** Baseline characteristics by residual cancer burden class. Values are n (% of the column class); the Total column gives n (% of the cohort). P values are from Monte-Carlo simulated chi-square tests (10,000 replicates), computed on observed categories only, because several cells are sparse.

| Characteristic | Total (n=328) | RCB-0 (n=90) | RCB-I (n=34) | RCB-II (n=143) | RCB-III (n=61) | P |
| --- | --- | --- | --- | --- | --- | --- |
| Age group |  |  |  |  |  | 0.084 |
| Young adult | 4 (1.2) | 0 (0.0) | 0 (0.0) | 4 (2.8) | 0 (0.0) |  |
| Early middle age | 37 (11.3) | 11 (12.2) | 5 (14.7) | 16 (11.2) | 5 (8.2) |  |
| Middle age | 101 (30.8) | 23 (25.6) | 10 (29.4) | 45 (31.5) | 23 (37.7) |  |
| Late middle age | 96 (29.3) | 32 (35.6) | 10 (29.4) | 45 (31.5) | 9 (14.8) |  |
| Older | 60 (18.3) | 13 (14.4) | 7 (20.6) | 26 (18.2) | 14 (23.0) |  |
| Very old | 30 (9.1) | 11 (12.2) | 2 (5.9) | 7 (4.9) | 10 (16.4) |  |
| Clinical stage before NAC |  |  |  |  |  | <0.001 |
| IA | 57 (17.4) | 16 (17.8) | 9 (26.5) | 24 (16.8) | 8 (13.1) |  |
| IB | 17 (5.2) | 6 (6.7) | 1 (2.9) | 9 (6.3) | 1 (1.6) |  |
| IIA | 102 (31.1) | 39 (43.3) | 9 (26.5) | 44 (30.8) | 10 (16.4) |  |
| IIB | 100 (30.5) | 21 (23.3) | 9 (26.5) | 50 (35.0) | 20 (32.8) |  |
| IIIA | 33 (10.1) | 4 (4.4) | 5 (14.7) | 6 (4.2) | 18 (29.5) |  |
| IIIB | 7 (2.1) | 0 (0.0) | 0 (0.0) | 5 (3.5) | 2 (3.3) |  |
| IIIC | 2 (0.6) | 2 (2.2) | 0 (0.0) | 0 (0.0) | 0 (0.0) |  |
| IV | 5 (1.5) | 1 (1.1) | 0 (0.0) | 4 (2.8) | 0 (0.0) |  |
| Unknown | 5 (1.5) | 1 (1.1) | 1 (2.9) | 1 (0.7) | 2 (3.3) |  |
| Molecular subtype |  |  |  |  |  | <0.001 |
| Luminal A | 47 (14.3) | 0 (0.0) | 4 (11.8) | 33 (23.1) | 10 (16.4) |  |
| Luminal B (HER2-negative) | 98 (29.9) | 20 (22.2) | 11 (32.4) | 49 (34.3) | 18 (29.5) |  |
| Luminal B (HER2-positive) | 63 (19.2) | 25 (27.8) | 6 (17.6) | 25 (17.5) | 7 (11.5) |  |
| HER2-enriched | 42 (12.8) | 22 (24.4) | 4 (11.8) | 11 (7.7) | 5 (8.2) |  |
| HER2-low | 21 (6.4) | 1 (1.1) | 2 (5.9) | 9 (6.3) | 9 (14.8) |  |
| Triple-negative | 37 (11.3) | 15 (16.7) | 6 (17.6) | 10 (7.0) | 6 (9.8) |  |
| Unknown | 20 (6.1) | 7 (7.8) | 1 (2.9) | 6 (4.2) | 6 (9.8) |  |
| ER |  |  |  |  |  | <0.001 |
| Negative | 82 (25.0) | 39 (43.3) | 10 (29.4) | 22 (15.4) | 11 (18.0) |  |
| Weakly positive | 39 (11.9) | 18 (20.0) | 4 (11.8) | 13 (9.1) | 4 (6.6) |  |
| Positive | 4 (1.2) | 2 (2.2) | 0 (0.0) | 2 (1.4) | 0 (0.0) |  |
| Strongly positive | 196 (59.8) | 27 (30.0) | 19 (55.9) | 104 (72.7) | 46 (75.4) |  |
| Unknown | 7 (2.1) | 4 (4.4) | 1 (2.9) | 2 (1.4) | 0 (0.0) |  |
| PR |  |  |  |  |  | <0.001 |
| Negative | 137 (41.8) | 58 (64.4) | 16 (47.1) | 45 (31.5) | 18 (29.5) |  |
| Weakly positive | 102 (31.1) | 18 (20.0) | 10 (29.4) | 50 (35.0) | 24 (39.3) |  |
| Positive | 2 (0.6) | 1 (1.1) | 0 (0.0) | 1 (0.7) | 0 (0.0) |  |
| Strongly positive | 80 (24.4) | 8 (8.9) | 7 (20.6) | 46 (32.2) | 19 (31.1) |  |
| Unknown | 7 (2.1) | 5 (5.6) | 1 (2.9) | 1 (0.7) | 0 (0.0) |  |
| HER2 |  |  |  |  |  | <0.001 |
| Negative | 197 (60.1) | 38 (42.2) | 22 (64.7) | 97 (67.8) | 40 (65.6) |  |
| HER2-low | 24 (7.3) | 3 (3.3) | 2 (5.9) | 10 (7.0) | 9 (14.8) |  |
| Equivocal | 12 (3.7) | 4 (4.4) | 0 (0.0) | 5 (3.5) | 3 (4.9) |  |
| Positive | 88 (26.8) | 40 (44.4) | 10 (29.4) | 29 (20.3) | 9 (14.8) |  |
| Unknown | 7 (2.1) | 5 (5.6) | 0 (0.0) | 2 (1.4) | 0 (0.0) |  |
| Ki-67 |  |  |  |  |  | <0.001 |
| Low | 71 (21.6) | 3 (3.3) | 7 (20.6) | 44 (30.8) | 17 (27.9) |  |
| Intermediate | 68 (20.7) | 18 (20.0) | 8 (23.5) | 30 (21.0) | 12 (19.7) |  |
| High | 171 (52.1) | 62 (68.9) | 18 (52.9) | 64 (44.8) | 27 (44.3) |  |
| Unknown | 18 (5.5) | 7 (7.8) | 1 (2.9) | 5 (3.5) | 5 (8.2) |  |
| Histologic grade |  |  |  |  |  | <0.001 |
| G1 | 19 (5.8) | 1 (1.1) | 1 (2.9) | 10 (7.0) | 7 (11.5) |  |
| G2 | 164 (50.0) | 29 (32.2) | 22 (64.7) | 84 (58.7) | 29 (47.5) |  |
| G3 | 132 (40.2) | 51 (56.7) | 10 (29.4) | 47 (32.9) | 24 (39.3) |  |
| Unknown | 13 (4.0) | 9 (10.0) | 1 (2.9) | 2 (1.4) | 1 (1.6) |  |
| Nodal / distant metastasis at diagnosis |  |  |  |  |  | <0.001 |
| Absent | 107 (32.6) | 53 (58.9) | 13 (38.2) | 35 (24.5) | 6 (9.8) |  |
| Present | 178 (54.3) | 18 (20.0) | 16 (47.1) | 97 (67.8) | 47 (77.0) |  |
| Unknown | 43 (13.1) | 19 (21.1) | 5 (14.7) | 11 (7.7) | 8 (13.1) |  |
| TILs |  |  |  |  |  | 0.093 |
| <10% | 179 (54.6) | 36 (40.0) | 19 (55.9) | 86 (60.1) | 38 (62.3) |  |
| 10-50% | 104 (31.7) | 34 (37.8) | 10 (29.4) | 45 (31.5) | 15 (24.6) |  |
| >50% | 25 (7.6) | 11 (12.2) | 3 (8.8) | 8 (5.6) | 3 (4.9) |  |
| Unknown | 20 (6.1) | 9 (10.0) | 2 (5.9) | 4 (2.8) | 5 (8.2) |  |

Eleven configurations were each evaluated across three algorithms and two SMOTE states (66 candidates; Supplementary Table S1). Among single-block models the radiologic block gave the highest test AUC (0.838) and the oncologic block was the strongest non-radiologic one (0.700), while the rest were individually of limited value (0.492–0.586) and the biochemical block alone fell below chance. Combinations without radiology ranged from 0.636 to 0.700, with no gain from adding blocks. Fig. 2 places the two metrics side by side and shows that the ranking of the leading configurations can change with the metric.

**Fig. 2.**
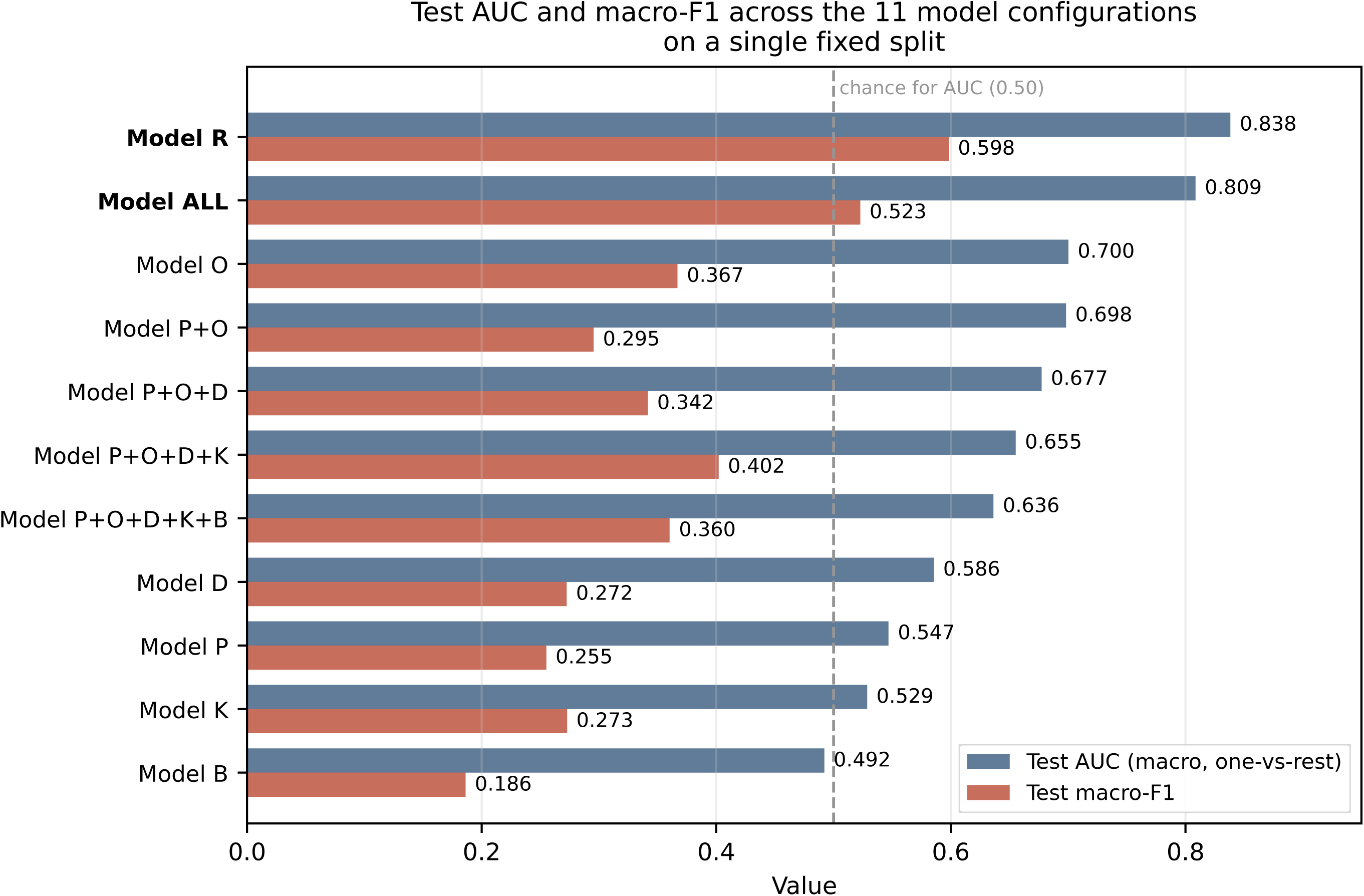
Test AUC (macro, one-vs-rest) and macro-F1 across the 11 evaluated model configurations, from a single fixed split. For each configuration the values shown are those of the best algorithm-SMOTE combination selected by inner cross-validation. The dashed line marks chance level for AUC (0.50); the biochemical block alone falls below it.

The ablation analysis showed which blocks earn their place (Supplementary Table S2). Removing the radiologic block lowered AUC by a mean paired difference of 0.170 (median 0.827 to 0.653; *d_z_*=5.46; p<0.001); removing the oncologic (0.033; *d_z_*=1.39) and pathologic (0.027; *d_z_*=1.34) blocks produced smaller but significant falls. Removing the demographic, comorbidity or biochemical blocks produced no measurable change (p=0.39, 0.70 and 0.90). The gain from multimodal integration therefore rests not on the number of blocks but on the combination of three specific ones.

### 3.2. Effect of the selection criterion

The two inner-CV criteria frequently rank the same candidates differently. Selecting on discrimination alone produced models that classified no patient correctly in RCB-III in 8 of 30 splits; with the mean of the two criteria this fell to 1 of 30, median RCB-III F1 rose from 0.154 to 0.286 and median macro-F1 from 0.564 to 0.601, at a cost of 0.020 in median AUC. What matters is the criterion itself rather than the tuning of its weights (Supplementary Table S7).

The stability of the selection differs between the models (Supplementary Table S8). For Model R, Random Forest without resampling was selected in 79% of splits; for Model ALL the selection was more dispersed (Random Forest with SMOTE 39%, XGBoost 35%, LightGBM 17%). Whichever was selected, the median external test AUC remained between 0.785 and 0.851, so selection uncertainty did not translate into performance uncertainty.

### 3.3. Performance across 100 repeated splits

The median test AUC was 0.823 (IQR 0.803–0.838) for Model ALL and 0.773 (0.746–0.803) for Model R (Table 2; Fig. 3), with Model ALL superior in 88% of splits (mean difference +0.050; *d_z_*=1.18; p=3.5×10⁻¹⁵), in 84% for macro-F1 (+0.069; *d_z_*=1.03) and 73% for accuracy (+0.051; *d_z_*=0.72). The gap between inner cross-validation and the held-out test ran at a median of 0.025 for Model ALL and 0.039 for Model R.

**Fig. 3.**
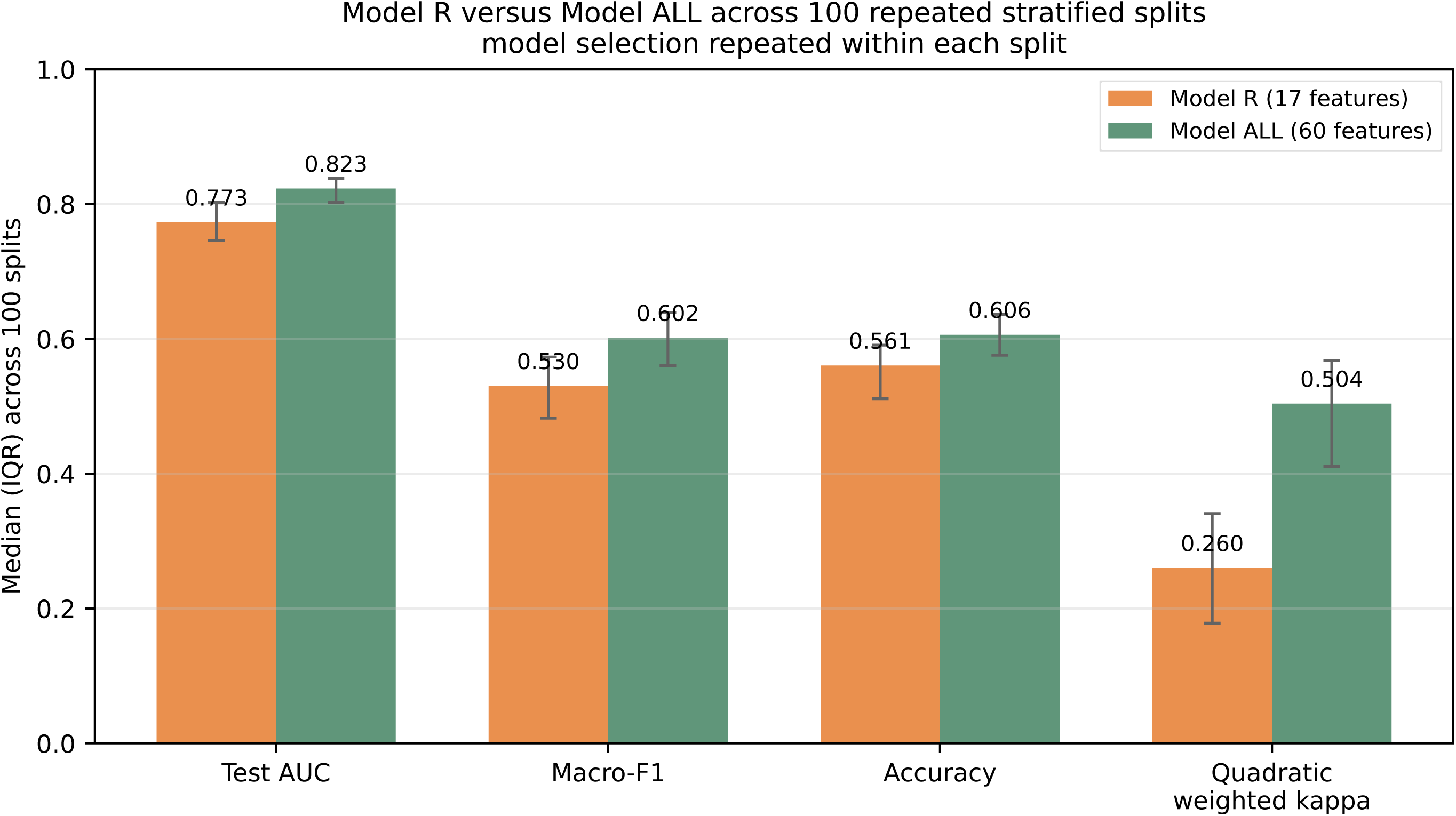
Performance of Model R and Model ALL across 100 stratified splits; bars show medians and error bars the interquartile range. Model selection was repeated within each split, using that split’s training data only. The largest difference is in ordinal agreement.

**Fig. 4.**
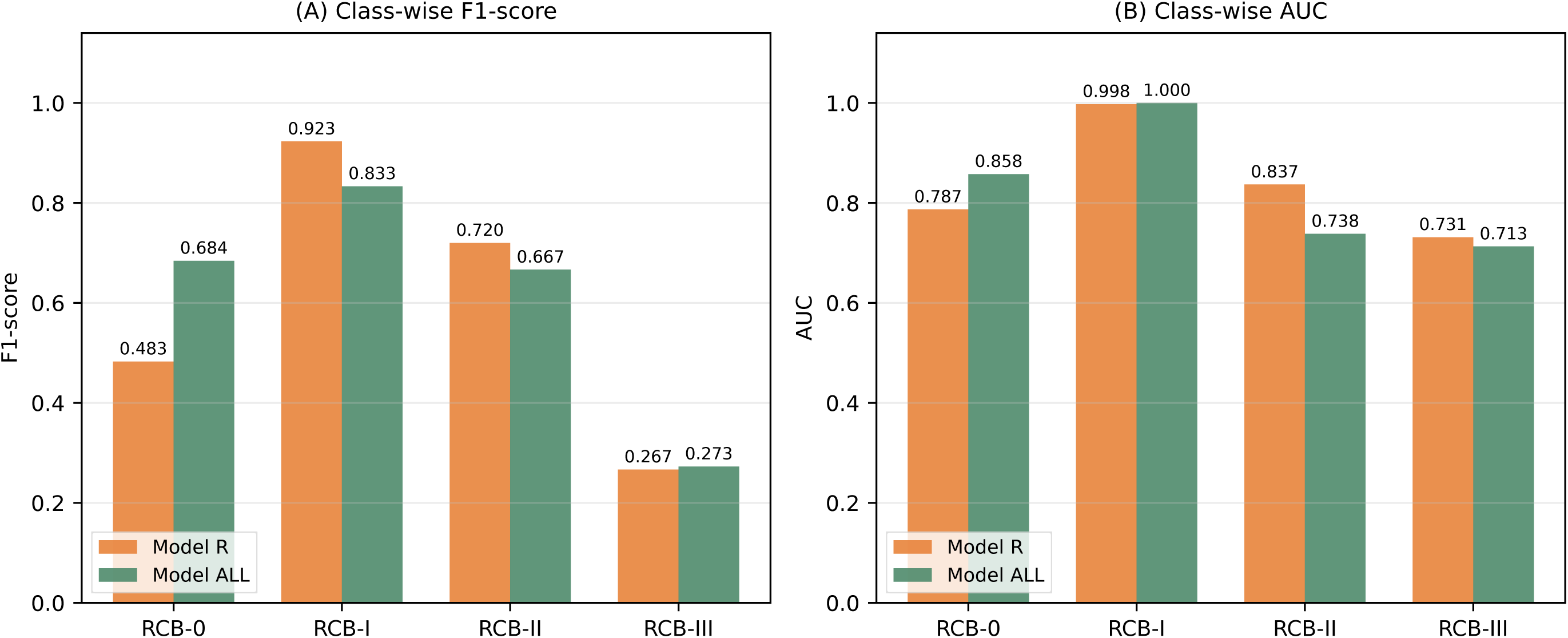
Class-wise comparison across RCB categories for the most frequently selected configuration: (A) F1-score and (B) AUC.

**Table 2.**
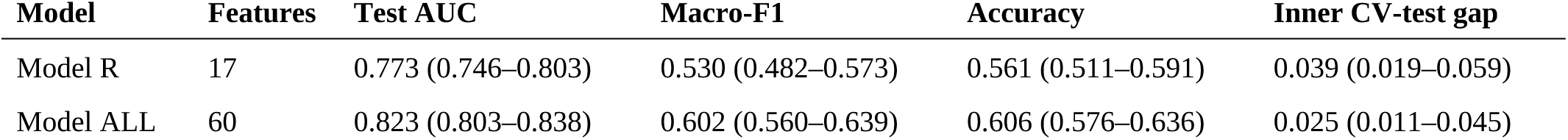
Median (IQR) performance across 100 stratified splits. Model selection was repeated within each split, using that split’s training data only.

| Model | Features | Test AUC | Macro-F1 | Accuracy | Inner CV-test gap |
| --- | --- | --- | --- | --- | --- |
| Model R | 17 | 0.773 (0.746–0.803) | 0.530 (0.482–0.573) | 0.561 (0.511–0.591) | 0.039 (0.019–0.059) |
| Model ALL | 60 | 0.823 (0.803–0.838) | 0.602 (0.560–0.639) | 0.606 (0.576–0.636) | 0.025 (0.011–0.045) |

### 3.4. Ordinal agreement and class-wise behaviour

The largest difference between the models lies in ordinal agreement (Table 3). The median quadratic weighted kappa was 0.504 (IQR 0.411–0.568) for Model ALL and 0.260 (0.178– 0.341) for Model R, with Model ALL superior in 98% of splits (mean difference +0.240; *d_z_*=1.69; p=8.3×10⁻¹⁸); adjacent-class accuracy rose from 0.742 to 0.811 and mean absolute error fell from 0.727 to 0.614. Because the clinical consequence of an adjacent-class error differs from that of a distant-class error, this difference is directly meaningful.

**Table 3.** Ordinal agreement metrics and performance at two clinically defined boundaries; medians across 100 stratified splits, with IQR in parentheses for the ordinal metrics. Thresholds were selected on training data only.

| **Metric (median across 100 splits)** | **Model R** | **Model ALL** |
| --- | --- | --- |
| Quadratic weighted kappa | 0.260 (0.178–0.341) | 0.504 (0.411–0.568) |
| Linearly weighted kappa | 0.282 (0.208–0.343) | 0.448 (0.385–0.510) |
| Adjacent-class accuracy | 0.742 (0.708–0.773) | 0.811 (0.788–0.848) |
| Mean absolute error | 0.727 (0.667–0.822) | 0.614 (0.557–0.697) |
| De-escalation boundary - AUC | 0.798 | 0.901 |
| De-escalation boundary - sensitivity / specificity (threshold 0.50) | 0.480 / 0.951 | 0.720 / 0.902 |
| De-escalation boundary - sensitivity / specificity (threshold from training) | 0.600 / 0.829 | 0.840 / 0.829 |
| Intensification boundary - AUC | 0.740 | 0.743 |
| Intensification boundary - sensitivity / specificity (threshold 0.50) | 0.000 / 1.000 | 0.083 / 0.981 |
| Intensification boundary - sensitivity / specificity (threshold from training) | 0.667 / 0.667 | 0.833 / 0.537 |

The multimodal model also leads in the highest-risk class: median RCB-III F1 0.333 versus 0.238, with the class lost entirely in 2 of 100 splits versus 5. On the single configuration shown in Fig. 3, class-wise F1 was 0.684 for RCB-0 (AUC 0.858; 95% CI 0.749–0.938), 0.833 for RCB-I (1.000, reflecting n=7 in the test set), 0.667 for RCB-II (0.738) and 0.273 for RCB-III (0.713). On that single split Model R retains a higher class-wise AUC in RCB-II (0.837) and RCB-III (0.731), a reversal not seen in the 100-split medians and consistent with the sampling variability documented in Section 3.5 (Supplementary Table S3).

### 3.5. What a single split would have shown

We examined, in our own data, what a single split used as the primary analysis would have shown. On the split generated with a fixed seed, the test AUC of Model R was 0.838, at the 98th percentile of its own distribution, and that of Model ALL 0.809, at the 31st; in macro-F1 the percentiles were 86 and 14. There the multimodal model appears inferior, although it is superior in 88% of splits. Inner cross-validation selected XGBoost there, whose median AUC across the 100 splits is 0.812, so it is the split rather than the selection that is unusual.

### 3.6. Decision threshold, calibration and validation

The two clinical boundaries behaved very differently. At de-escalation the multimodal model is strong and clearly ahead: median AUC 0.901 versus 0.798, sensitivity 0.720 versus 0.480 at the default threshold, net benefit 0.212, and sensitivity never zero. At intensification the two models are almost identical (median AUC 0.743 versus 0.740) and the default 0.50 threshold collapses for both, marking no patient positive in 48 of 100 splits for Model ALL and 51 for Model R. This is a structural problem of default arg-max assignment in an imbalanced multiclass outcome, not a defect of one model.

Selecting the threshold by cross-validation on training data alone (Youden J, median 0.22) raised the median sensitivity at intensification from 0.083 to 0.833, lowered specificity from 0.981 to 0.537, raised net benefit from 0.000 to 0.047 and left no split with zero sensitivity (Supplementary Table S6; Supplementary Fig. S17). At de-escalation it raised sensitivity from 0.720 to 0.840 and lowered specificity to 0.829; for the radiology model it raised intensification sensitivity from 0.000 to 0.667. High sensitivity there costs flagging roughly half of all patients unnecessarily, an operating point that suits an alert layer rather than a definitive decision rule.

In the 5×5 two-level resampling the inner-loop AUC averaged 0.815±0.015, the outer-test AUC 0.825±0.040 and the mean difference −0.010±0.053 (Supplementary Table S4; Supplementary Fig. S6); the inner value sitting slightly below the outer one is expected, since inner folds train on a smaller subset. The mean Brier score of the probability outputs was 0.132, moved to 0.123 by Platt scaling and 0.127 by isotonic regression (Supplementary Fig. S15); the gain is small and the reported model uses uncalibrated probabilities.

Two sensitivity analyses were conducted across 30 paired splits each. Excluding the treatment-process variables moved the median test AUC from 0.818 to 0.827 and macro-F1 from 0.601 to 0.617, and reduced the splits in which RCB-III was lost from 1 to 0. Imputing before the split lowered the median test AUC of Model ALL by 0.002 and left Model R unaffected (*d_z_*=-0.13; p=0.529).

In the global SHAP ranking the most influential variables were metastasis site, BI-RADS, calcification distribution, mass density, calcification morphology, prior surgery, lesion type and metastasis status; six of the top eight are radiologic and two oncologic, so this ranking coincides with the ablation analysis (Supplementary Figs. S7–S11).

## 4. Discussion

This study set out to predict four-class residual cancer burden from pre-treatment data and found that model selection itself became a finding. The multimodal model discriminated better in 88% of 100 repeated splits, and the difference is larger still in ordinal agreement, where it holds in 98%: the model errs closer to the true class when it errs.

The metric-dependent divergence recurs at several scales: the six candidates of a single model rank differently under the two inner-CV criteria, the AUC and macro-F1 orderings diverge across configurations, and selecting on discrimination alone lost the RCB-III class entirely in 8 of 30 splits. Selection on a single discrimination metric can therefore prefer a model that writes off the clinically most critical class.

The decision rule requires separate design as well. The default arg-max assignment marked no patient as positive at the intensification boundary in 48 of 100 splits, and in 51 for the radiology model, so the problem is structural rather than model-specific; once the threshold is selected from training data alone, the same models give median sensitivities of 0.833 and 0.667 there. Many published multiclass models report low sensitivity in sparse classes as a limitation of the model without treating threshold selection as a separate design decision.

We applied our argument about evaluation design to our own analysis: resting the primary analysis on one split would have contradicted the principle the study defends, so it was moved to 100 splits with selection repeated in each. We also report what one fixed split would have shown: there Model R fell at the 98th percentile of its distribution and Model ALL at the 31st, and the multimodal model appeared inferior. This is not an abstract caution but an instance measured in the same dataset [15–18].

The block-structured design interrogates the source of the advantage from two directions: cumulative addition of blocks reads it forward, ablation of each block in turn reads it backward. Either alone is exposed to sampling chance; when they agree, the attribution is strengthened. The demographic, comorbidity and biochemical blocks that brought no gain on addition are those whose removal costs nothing, whereas removing the radiologic block lowers AUC by 0.170, and the global SHAP ranking points to the same blocks. Multimodal integration therefore rests not on the number of blocks but on three specific ones, which answers concretely which data layers are worth collecting [9,10].

The timing of the predictors is a deliberate design decision. Treatment-process variables carry reverse causality in an observational cohort and invite a causal reading that an observational model cannot support; that excluding them did not lower performance indicates the model does not lean on this signal.

The evaluation of class-imbalance interventions is likewise criterion-dependent. SMOTE did not improve discrimination but raised macro-F1 and ordinal agreement, and it appears in the most frequently selected configuration. Imbalance corrections have been shown not to improve discrimination, to be capable of degrading calibration, and to bring gains obtainable by shifting the probability threshold instead [30,31]. Our data show both mechanisms at work, and calibration was not degraded (raw Brier 0.132).

Positioned as a pre-treatment rather than a preoperative tool, the model occupies a different clinical place. At de-escalation the median AUC is 0.901 and net benefit is positive across the 0.05–0.80 threshold range, so flagging patients with a high probability of complete response before treatment begins appears feasible. At intensification high sensitivity is achievable but costs flagging roughly half of all patients unnecessarily; there the model is better positioned as an alert layer than as a decision instrument.

The outcome definition also departs from common practice. Prediction studies narrow the four-tier scale almost without exception [12–14], hiding the two boundaries at which adjuvant intensity is decided. The narrowing has a statistical rationale, and that cost is visible here, with the sparsest class represented by 12 test patients. Retaining four classes nonetheless allowed ordinal agreement and the two clinical boundaries to be evaluated separately, neither obtainable within a binary framing.

### 4.1. Limitations

● Data were assembled retrospectively from a single centre; selection bias and centre-specific practice limit generalisability, and no external validation was performed. The cohort is exclusively female, so findings cannot be generalised to male breast cancer; no formal fairness analysis across sociodemographic subgroups was undertaken.
● Study size was not based on formal sample size calculations [32,33]; for a four-class outcome this constrains the effective sample, with RCB-I represented by 7 and RCB-III by 12 patients in the test set.
● The 100 splits are drawn from the same 328 patients and their training sets overlap substantially; the reported p values reflect split-level pairing and should not be read as a count of independent observations.
● Hyperparameters were held fixed with no configuration-specific tuning; while this focuses the comparison on block composition, both models may perform below their attainable best. Imputation preceded the split; its effect was not statistically detectable, but split-respecting imputation should be standard in future validation.
● Decision thresholds were learned within the same cohort, so their external validity is unknown and they would require re-estimation before any threshold-based clinical use.
● Analyses requiring a single model used the most frequently selected configuration, chosen in 39% of splits; explainability and calibration may differ where another configuration was selected.
● Four biochemical variables carried no variance and could not contribute, and radiologic variables were derived from structured reports, with raw images and radiomic features not included.

The model remains investigational. Priority steps are multicentre prospective external validation with pre-specified thresholds, cost-sensitive strategies for RCB-III sensitivity, and extension to radiomic imaging features.

## 5. Conclusions

For four-class RCB prediction from pre-treatment data, the multimodal model was superior to the radiology-focused model in discrimination in 88% of splits and in ordinal agreement in 98%, measured across 100 stratified splits with model selection repeated within each split.

The principal contribution is to show that model development for an imbalanced multiclass clinical outcome requires three separate design decisions. Evaluation resting on a single split can reverse the ranking of models; a selection criterion resting on discrimination alone can lose the highest-risk class entirely; and the default decision threshold can render an available signal unusable. All three are resolved by moving evaluation to repeated resampling and determining the criterion and the threshold separately, from training data alone.

## Acknowledgements and declarations

## Acknowledgements

None.

## Ethical approval

The study was approved by the Non-Interventional Clinical Research Ethics Committee of Dokuz Eylul University (approval no. 2024/25-08; date of approval 17 July 2024). Given the retrospective, non-interventional design, the committee waived the requirement for individual informed consent; all data were anonymised before analysis.

## Data availability

The anonymised dataset is available from the corresponding author upon reasonable request, subject to restrictions related to patient privacy.

## Patient and public involvement

No patients or members of the public were involved in the design, conduct, reporting or dissemination of this research.

## Code availability

The analysis code and result files are openly available in a transparency archive deposited on Zenodo with a persistent identifier (doi:10.5281/zenodo.23165375). The archive contains the repeated-split primary analysis, the selection-criterion and decision-threshold analyses, the ablation and two sensitivity analyses, the result files underlying every table in the manuscript, the fitted model object, the feature list and class order, all twenty figures in vector form, and an English README mapping each script to the reported figures and tables. Code is released under the MIT Licence, documentation and result files under CC BY 4.0.

Declaration of generative AI and AI-assisted technologies in the manuscript preparation process. During the preparation of this work the authors used Claude (Anthropic) in order to improve language and readability. The authors reviewed and edited the content as needed and take full responsibility for the content of the publication.

## References

[1] Loibl S, Poortmans P, Morrow M, Denkert C, Curigliano G. Breast cancer. Lancet. 2021;397(10286):1750–69. doi:10.1016/S0140-6736(20)32381-3

[2] Curigliano G, Burstein HJ, Gnant M, et al. Understanding breast cancer complexity to improve patient outcomes: the St Gallen International Consensus Conference 2023. Ann Oncol. 2023;34(11):970–86. doi:10.1016/j.annonc.2023.08.017

[3] Loibl S, André F, Bachelot T, et al. Early breast cancer: ESMO Clinical Practice Guideline for diagnosis, treatment and follow-up. Ann Oncol. 2024;35(2):159–82. doi:10.1016/j.annonc.2023.11.016

[4] Spring LM, Fell G, Arfe A, et al. Pathological complete response after neoadjuvant chemotherapy and impact on breast cancer recurrence and survival: a comprehensive meta-analysis. Clin Cancer Res. 2020;26(12):2838–48. doi:10.1158/1078-0432.CCR-19-3492

[5] Cortazar P, Zhang L, Untch M, et al. Pathological complete response and long-term clinical benefit in breast cancer: the CTNeoBC pooled analysis. Lancet. 2014;384(9938):164–72. doi:10.1016/S0140-6736(14)62422-8

[6] Symmans WF, Peintinger F, Hatzis C, et al. Measurement of residual breast cancer burden to predict survival after neoadjuvant chemotherapy. J Clin Oncol. 2007;25(28):4414–22. doi:10.1200/JCO.2007.10.6823

[7] Symmans WF, Wei C, Gould R, et al. Long-term prognostic risk after neoadjuvant chemotherapy associated with residual cancer burden and breast cancer subtype. J Clin Oncol. 2017;35(10):1049–60. doi:10.1200/JCO.2015.63.1010

[8] Yau C, Osdoit M, van der Noordaa M, et al. Residual cancer burden after neoadjuvant chemotherapy and long-term survival outcomes in breast cancer: a multicentre pooled analysis of 5161 patients. Lancet Oncol. 2022;23(1):149–60. doi:10.1016/S1470-2045(21)00589-1

[9] Acosta JN, Falcone GJ, Rajpurkar P, Topol EJ. Multimodal biomedical AI. Nat Med. 2022;28(9):1773–84. doi:10.1038/s41591-022-01981-2

[10] Kline A, Wang H, Li Y, et al. Multimodal machine learning in precision health: a scoping review. NPJ Digit Med. 2022;5(1):171. doi:10.1038/s41746-022-00712-8

[11] Cain EH, Saha A, Harowicz MR, Marks JR, Marcom PK, Mazurowski MA. Multivariate machine learning models for prediction of pathologic response to neoadjuvant therapy in breast cancer using MRI features. Breast Cancer Res Treat. 2019;173(2):455–63. doi:10.1007/s10549-018-4990-9

[12] Janse MHA, Janssen LM, van der Velden BHM, et al. Deep learning-based segmentation of locally advanced breast cancer on MRI in relation to residual cancer burden: a multiinstitutional cohort study. J Magn Reson Imaging. 2023;58(6):1739–49. doi:10.1002/jmri.28679

[13] Valderrama NF, Morel LO, Mweze DT, et al. Chemo-prAIdict Breast: a deep learning solution for predicting residual disease on biopsies of breast cancer patients treated with neoadjuvant chemotherapy. Eur J Cancer. 2026;234:116222. doi:10.1016/j.ejca.2026.116222

[14] Li W, Huang YH, Zhu T, et al. Noninvasive artificial intelligence system for early predicting residual cancer burden during neoadjuvant chemotherapy in breast cancer. Ann Surg. 2025;281(4):645–54. doi:10.1097/SLA.0000000000006279

[15] Andaur Navarro CL, Damen JAA, Takada T, et al. Risk of bias in studies on prediction models developed using supervised machine learning techniques: systematic review. BMJ. 2021;375:n2281. doi:10.1136/bmj.n2281

[16] Vabalas A, Gowen E, Poliakoff E, Casson AJ. Machine learning algorithm validation with a limited sample size. PLoS One. 2019;14(11):e0224365. doi:10.1371/journal.pone.0224365

[17] Varma S, Simon R. Bias in error estimation when using cross-validation for model selection. BMC Bioinformatics. 2006;7:91. doi:10.1186/1471-2105-7-91

[18] Krstajic D, Buturovic LJ, Leahy DE, Thomas S. Cross-validation pitfalls when selecting and assessing regression and classification models. J Cheminform. 2014;6:10. doi:10.1186/1758-2946-6-10

[19] Bates DW, Auerbach A, Schulam P, Wright A, Saria S. Reporting and implementing interventions involving machine learning and artificial intelligence. Ann Intern Med. 2020;172(11 Suppl):S137–44. doi:10.7326/M19-0872

[20] Lundberg SM, Erion G, Chen H, et al. From local explanations to global understanding with explainable AI for trees. Nat Mach Intell. 2020;2(1):56–67. doi:10.1038/s42256-019-0138-9

[21] Collins GS, Moons KGM, Dhiman P, et al. TRIPOD+AI statement: updated guidance for reporting clinical prediction models that use regression or machine learning methods. BMJ. 2024;385:e078378. doi:10.1136/bmj-2023-078378

[22] Breiman L. Random forests. Mach Learn. 2001;45(1):5–32. doi:10.1023/A:1010933404324

[23] Chen T, Guestrin C. XGBoost: a scalable tree boosting system. In: Proc 22nd ACM SIGKDD Int Conf Knowledge Discovery and Data Mining. New York: ACM; 2016. p. 785–94. doi:10.1145/2939672.2939785

[24] Ke G, Meng Q, Finley T, et al. LightGBM: a highly efficient gradient boosting decision tree. In: Advances in Neural Information Processing Systems 30. Red Hook (NY): Curran Associates; 2017. p. 3146–54.

[25] Chawla NV, Bowyer KW, Hall LO, Kegelmeyer WP. SMOTE: synthetic minority over-sampling technique. J Artif Intell Res. 2002;16:321–57. doi:10.1613/jair.953

[26] Lundberg SM, Lee SI. A unified approach to interpreting model predictions. In: Advances in Neural Information Processing Systems 30. Red Hook (NY): Curran Associates; 2017. p. 4765–74.

[27] Van Calster B, McLernon DJ, van Smeden M, Wynants L, Steyerberg EW. Calibration: the Achilles heel of predictive analytics. BMC Med. 2019;17(1):230. doi:10.1186/s12916-019-1466-7

[28] Vickers AJ, Elkin EB. Decision curve analysis: a novel method for evaluating prediction models. Med Decis Making. 2006;26(6):565–74. doi:10.1177/0272989X06295361

[29] Vickers AJ, van Calster B, Steyerberg EW. A simple, step-by-step guide to interpreting decision curve analysis. Diagn Progn Res. 2019;3:18. doi:10.1186/s41512-019-0064-7

[30] van den Goorbergh R, van Smeden M, Timmerman D, Van Calster B. The harm of class imbalance corrections for risk prediction models: illustration and simulation using logistic regression. J Am Med Inform Assoc. 2022;29(9):1525–34. doi:10.1093/jamia/ocac093

[31] Carriero A, Luijken K, de Hond A, Moons KGM, van Calster B, van Smeden M. The harms of class imbalance corrections for machine learning based prediction models: a simulation study. Stat Med. 2025;44(3-4):e10320. doi:10.1002/sim.10320

[32] Riley RD, Ensor J, Snell KIE, et al. Calculating the sample size required for developing a clinical prediction model. BMJ. 2020;368:m441. doi:10.1136/bmj.m441

[33] Pate A, Riley RD, Collins GS, et al. Minimum sample size for developing a multivariable prediction model using multinomial logistic regression. Stat Methods Med Res. 2023;32(3):555–71. doi:10.1177/09622802231151220

