## Supplementary Material (Tables S1-S8 and Figures S1-S17 for "Predicting Four-Class Residual Cancer Burden (RCB) Before Neoadjuvant Chemotherapy in Breast Cancer: Block-Structured Model Comparison, Selection Criterion and Decision Threshold"

### Supplementary Table S1. Best configuration per model on a single split

Best algorithm-SMOTE combination for each of the 11 modular configurations, chosen from 66 candidates on a single fixed split. Selection used only the training data, ranking candidates by the mean of macro AUC and macro-F1 obtained in five-fold inner cross-validation. This table shows how the configurations appear on one split; the primary analysis rests on 100 splits (Table 2).

| Model | Features | Algorithm | SMOTE | Inner AUC | Inner F1 | Selection score | Test AUC | Macro-F1 | Accuracy |
|---|---|---|---|---|---|---|---|---|---|
| Model R | 17 | Random Forest | No | 0.739 | 0.499 | 0.619 | 0.838 | 0.598 | 0.636 |
| Model ALL | 60 | XGBoost | No | 0.830 | 0.641 | 0.735 | 0.809 | 0.523 | 0.591 |
| Model O | 3 | Random Forest | No | 0.597 | 0.373 | 0.485 | 0.700 | 0.367 | 0.515 |
| Model P+O | 14 | Random Forest | No | 0.659 | 0.382 | 0.520 | 0.698 | 0.295 | 0.500 |
| Model P+O+D | 19 | Random Forest | Yes | 0.656 | 0.360 | 0.508 | 0.677 | 0.342 | 0.409 |
| Model P+O+D+K | 29 | XGBoost | Yes | 0.682 | 0.436 | 0.559 | 0.655 | 0.402 | 0.500 |
| Model P+O+D+K+B | 43 | XGBoost | Yes | 0.677 | 0.411 | 0.544 | 0.636 | 0.360 | 0.500 |
| Model D | 5 | LightGBM | Yes | 0.508 | 0.290 | 0.399 | 0.586 | 0.272 | 0.303 |
| Model P | 11 | Random Forest | No | 0.622 | 0.314 | 0.468 | 0.547 | 0.255 | 0.424 |
| Model K | 10 | Random Forest | No | 0.520 | 0.260 | 0.390 | 0.529 | 0.273 | 0.470 |
| Model B | 14 | LightGBM | Yes | 0.516 | 0.238 | 0.377 | 0.492 | 0.186 | 0.258 |

### Supplementary Table S2. Ablation analysis - block removal

Each block was removed from Model ALL in turn and the paired change in test AUC measured across 20 stratified splits. The difference column is the mean of the paired differences across splits, not the difference of medians; a positive value means that removing the block lowered performance, that is, the block contributes. The median AUC of the full model is 0.827. P values are from the paired Wilcoxon signed-rank test.

| Block removed | Features | Remaining model AUC (median) | Mean paired difference | $d_z$ | p |
|---|---|---|---|---|---|
| Radiologic | 17 | 0.653 | +0.170 | 5.46 | <0.001 |
| Oncologic | 3 | 0.789 | +0.033 | 1.39 | <0.001 |
| Pathologic | 11 | 0.800 | +0.027 | 1.34 | <0.001 |
| Demographic | 5 | 0.818 | +0.004 | 0.23 | 0.388 |
| Comorbidity | 10 | 0.823 | +0.001 | 0.06 | 0.701 |
| Biochemical | 14 | 0.826 | -0.001 | -0.07 | 0.898 |

Three blocks make a measurable contribution and three do not. This ordering coincides with the global SHAP ranking, in which six of the top eight variables are radiologic and two oncologic.

### Supplementary Table S3. Class-wise test performance

Class-wise metrics for the most frequently selected configuration (Random Forest with SMOTE) on the held-out test set (n=66), with bootstrap 95% confidence intervals for AUC (B=500). The mean of the class-wise F1 values equals the macro-F1 (0.614) and the mean of the class-wise AUC values the macro AUC (0.827). The perfect AUC for RCB-I reflects the sparsity of that class in the test set (n=7).

| RCB class | Precision | Sensitivity | F1 | AUC | AUC 95% CI | Specificity |
|---|---|---|---|---|---|---|
| RCB-0 | 0.650 | 0.722 | 0.684 | 0.858 | 0.749–0.938 | 0.854 |
| RCB-I | 1.000 | 0.714 | 0.833 | 1.000 | 1.000–1.000 | 1.000 |
| RCB-II | 0.645 | 0.690 | 0.667 | 0.738 | 0.621–0.845 | 0.703 |
| RCB-III | 0.300 | 0.250 | 0.273 | 0.713 | 0.553–0.862 | 0.870 |

### Supplementary Table S4. Two-level nested resampling

5×5 nested resampling for the most frequently selected configuration. Because no model selection occurs in the inner loop, this is a stability check comparing inner- and outer-loop performance rather than nested cross-validation in the classical sense. The inner value sitting slightly below the outer one is expected, since inner folds train on a smaller subset.

| Metric | Value |
|---|---|
| Inner-loop AUC (mean ± SD) | 0.815 ± 0.015 |
| Outer test AUC (mean ± SD) | 0.825 ± 0.040 |
| Mean difference (inner - outer) | -0.010 ± 0.053 |
| Number of outer folds | 25 |

### Supplementary Table S5. Paired comparison across 100 stratified splits

Model selection was repeated within each split using that split's training data only, and the same split was given to both models. P values are from the paired Wilcoxon signed-rank test and effect sizes from Cohen's $d_z$. Because the splits are drawn from the same 328 patients, these p values should not be read as a count of independent observations.

| Metric | Splits favouring ALL | Mean difference | $d_z$ | p |
|---|---|---|---|---|
| Test AUC | 88% | +0.050 | 1.18 | $3.5\times10^{-15}$ |
| Macro-F1 | 84% | +0.069 | 1.03 | $2.0\times10^{-13}$ |
| Accuracy | 73% | +0.051 | 0.72 | $1.9\times10^{-9}$ |
| Quadratic weighted kappa | 98% | +0.240 | 1.69 | $8.3\times10^{-18}$ |
| Adjacent-class accuracy | 83% | +0.074 | 1.27 | $3.6\times10^{-15}$ |

The largest and most consistent difference is in ordinal agreement. In the RCB-III class the median F1 was 0.333 for Model ALL and 0.238 for Model R, with the class lost entirely in 2 of 100 and 5 of 100 splits respectively.

### Supplementary Table S6. Decision threshold rules

Performance of three threshold rules at the two clinical boundaries across 100 stratified splits. Thresholds were selected by cross-validation on the training data only and evaluated on the test set. Values are median (IQR).

| Boundary | Threshold rule | Median threshold | Sensitivity | Specificity | Net benefit |
|---|---|---|---|---|---|
| De-escalation | Default 0.50 | 0.50 | 0.720 (0.640–0.760) | 0.902 (0.878–0.927) | +0.212 |
| De-escalation | Youden J (training) | 0.40 | 0.840 (0.760–0.880) | 0.829 (0.805–0.878) | +0.243 |
| De-escalation | Lowest threshold with training sensitivity ≥ 0.50 | 0.60 | 0.560 (0.520–0.640) | 0.951 (0.927–0.976) | +0.160 |
| Intensification | Default 0.50 | 0.50 | 0.083 (0.000–0.083) | 0.981 (0.963–1.000) | +0.000 |
| Intensification | Youden J (training) | 0.22 | 0.833 (0.729–0.917) | 0.537 (0.458–0.611) | +0.047 |
| Intensification | Lowest threshold with training sensitivity ≥ 0.50 | 0.32 | 0.500 (0.417–0.667) | 0.750 (0.704–0.796) | -0.006 |

At the intensification boundary, sensitivity was zero in 48 of 100 splits with the default threshold and in none with thresholds selected from training data. No rule produced zero sensitivity at the de-escalation boundary. The third rule selects the highest threshold at which sensitivity in the training folds still reaches 0.50; it is not recommended at the intensification boundary, where its net benefit on the test partitions falls below zero.

### Supplementary Table S7. Comparison of selection criteria

Behaviour of three selection rules across 30 stratified splits. In all rules selection used the training data only; the sole difference is the metric used in inner cross-validation.

| Selection rule | Most frequent choice | Median test AUC | Median macro-F1 | Median RCB-III F1 | RCB-III F1 = [illegible] |
|---|---|---|---|---|---|
| K1 inner-CV AUC | Random Forest / No (93%) | 0.838 | 0.564 | 0.154 | 8/30 |
| K2 inner-CV macro-F1 | Random Forest / Yes (50%) | 0.817 | 0.599 | 0.291 | 1/30 |
| K3 mean of the two | Random Forest / Yes (53%) | 0.818 | 0.601 | 0.286 | 1/30 |

The main analysis used K3. What matters is the criterion itself rather than the tuning of its weights: K2 resolves the same problem but leaves the selection less stable.

### Supplementary Table S8. Selection frequency and external performance by selection

Distribution of the selection repeated within each of the 100 stratified splits. Whichever configuration is selected, the median external test AUC stays within a narrow band; selection uncertainty does not translate into performance uncertainty.

| Model | Selected configuration | n / 100 | Median test AUC | Median macro-F1 |
|---|---|---|---|---|
| Model R | Random Forest / no SMOTE | 79 | 0.786 | 0.533 |
| Model R | XGBoost / no SMOTE | 9 | 0.745 | 0.489 |
| Model R | LightGBM / no SMOTE | 5 | 0.739 | 0.518 |
| Model R | Random Forest / SMOTE | 5 | 0.761 | 0.527 |
| Model ALL | Random Forest / SMOTE | 39 | 0.828 | 0.611 |
| Model ALL | XGBoost / no SMOTE | 35 | 0.812 | 0.590 |
| Model ALL | LightGBM / no SMOTE | 17 | 0.820 | 0.595 |

| Model | Selected configuration | n / 100 | Median test AUC | Median macro-F1 |
|---|---|---|---|---|
| Model ALL | Random Forest / no SMOTE | 5 | 0.851 | 0.653 |
| Model ALL | XGBoost / SMOTE | 4 | 0.785 | 0.523 |

### Supplementary Table S9. Model blocks and their contents

Candidate variables organised into six thematic blocks. Four biochemical variables (ALT, AST, LDH, TSH) take a single value across the cohort and enter no split, so the effective feature count is 56 for Model ALL and 16 for Model R.

| Block / Model | Features | Content |
|---|---|---|
| Model P | 11 | Histologic type; ER; PR; HER2; molecular subtype; Ki-67; tubule grade; nuclear grade; mitotic grade; histologic grade; TILs |
| Model O | 3 | Metastasis status; metastasis site; clinical stage before NAC |
| Model D | 5 | Affected breast; BMI class; age group; blood type; sun exposure |
| Model K | 10 | Hypertension; diabetes; COPD; smoking; family history of breast cancer; thyroid disease; retinopathy; neuropathy; osteoporosis; depression |
| Model B | 14 (10 effective) | ALP; ALT; AST; BUN; CA 15-3; CEA; CRP; GGT; glucose; HbA1c; creatinine; LDH; TSH; eGFR |
| Model R | 17 | BI-RADS; breast density; localisation; lesion type; architectural distortion; mass shape; mass margin; mass density; calcification morphology; calcification distribution; asymmetry; multifocality; lesion stable for 2 years; skin retraction; nipple retraction; prior surgery; cosmetic implant |
| Model ALL | 60 (56 effective) | 11 pathologic + 3 oncologic + 5 demographic + 10 comorbidity + 14 biochemical + 17 radiologic |

### Supplementary Table S10. Hyperparameter values

All hyperparameters were fixed at these pre-specified values for every configuration and every split; no search was performed. Values not listed are the library defaults of the versions given in Section 2.7. SMOTE was applied with default settings (k_neighbors=5) within training folds only, and was skipped in any fold where the smallest class held fewer than six observations.

| Algorithm | Hyperparameter | Value |
|---|---|---|
| Random Forest | n_estimators | 200 |
| | max_depth | 10 |
| | min_samples_split | 5 |
| | min_samples_leaf | 2 |
| XGBoost | n_estimators | 200 |
| | max_depth | 6 |
| | learning_rate | 0.1 |
| | subsample | 0.8 |
| | colsample_bytree | 0.8 |

| Algorithm | Hyperparameter | Value |
|---|---|---|
| LightGBM | n_estimators | 200 |
| | max_depth | 6 |
| | learning_rate | 0.1 |
| | num_leaves | 31 |
| | subsample | 0.8 |
| | colsample_bytree | 0.8 |
| Imputation | Random Forest trees | 100 |
| All models | random_state | fixed per split (0–99) |

### Supplementary figure captions

Supplementary Fig. S1. Confusion matrix for the most frequently selected configuration on the held-out test set; 41 of 66 patients were classified correctly.

Supplementary Fig. S2. Class-wise receiver operating characteristic curves for Model ALL.

Supplementary Fig. S3. Class-wise precision-recall curves for Model ALL.

Supplementary Fig. S4. Class-wise cumulative gain curves for Model ALL.

Supplementary Fig. S5. Class-wise lift curves for Model ALL.

Supplementary Fig. S6. Difference between inner-CV AUC and outer test AUC across the 25 outer folds of the 5×5 nested resampling; mean -0.012.

Supplementary Fig. S7. Global feature importance for Model ALL, expressed as the mean absolute SHAP value across RCB classes.

Supplementary Fig. S8. Class-specific SHAP summary for RCB-0.

Supplementary Fig. S9. Class-specific SHAP summary for RCB-I.

Supplementary Fig. S10. Class-specific SHAP summary for RCB-II.

Supplementary Fig. S11. Class-specific SHAP summary for RCB-III.

Supplementary Fig. S12. Distribution of test AUC for Model R and Model ALL across 100 stratified splits, with model selection repeated within each split.

Supplementary Fig. S13. Distribution of the paired per-split difference in test AUC (Model ALL minus Model R) across the same 100 splits.

Supplementary Fig. S14. Distribution of quadratic weighted kappa across the same 100 splits; this is the metric on which the two models differ most.

Supplementary Fig. S15. Calibration curves for Model ALL, comparing raw probabilities, Platt scaling and isotonic regression. Mean Brier scores are 0.132, 0.123 and 0.127 respectively.

Supplementary Fig. S16. Decision curve analysis at the two clinical boundaries: de-escalation (RCB-0/I versus RCB-II/III) and intensification (RCB-III versus the rest). The dotted curve is the treat-all strategy and the horizontal line the treat-none strategy.

Supplementary Fig. S17. Decision threshold sweep. Sensitivity, specificity and F1 as a function of threshold at the two clinical boundaries; vertical lines mark the three threshold rules selected from the training data.

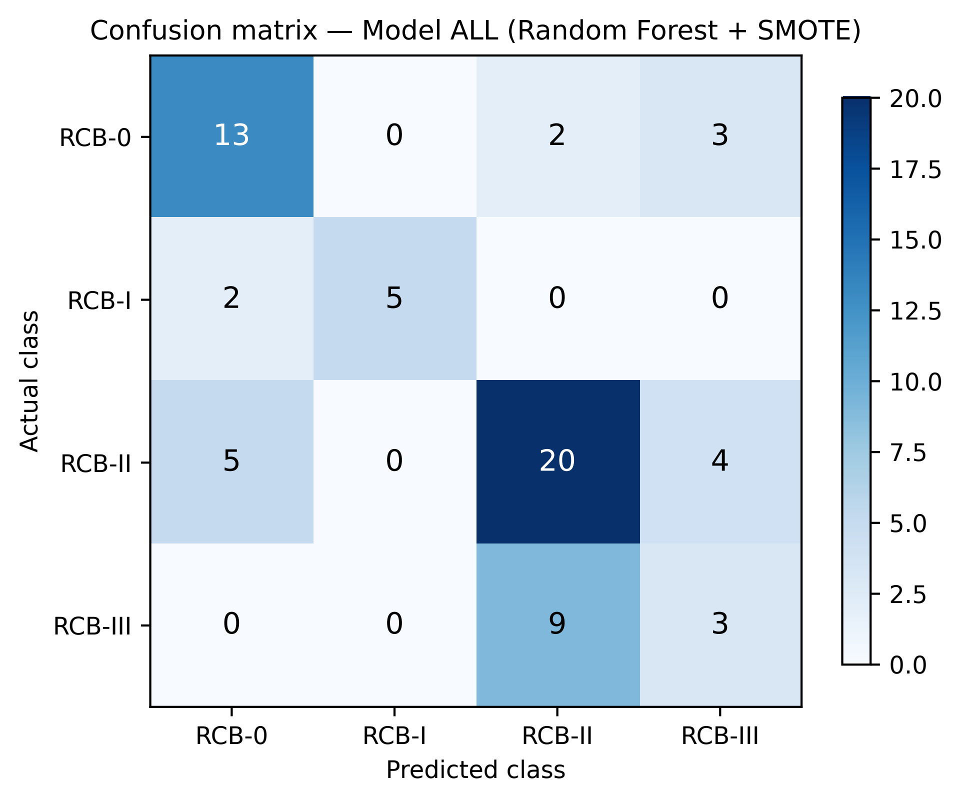

Confusion matrix — Model ALL (Random Forest + SMOTE)
RCB-0
RCB-I
RCB-II
RCB-III
13
0
2
3
2
5
0
0
5
0
20
4
0
0
9
3
Actual class
Predicted class
20.0
17.5
15.0
12.5
10.0
7.5
5.0
2.5
0.0

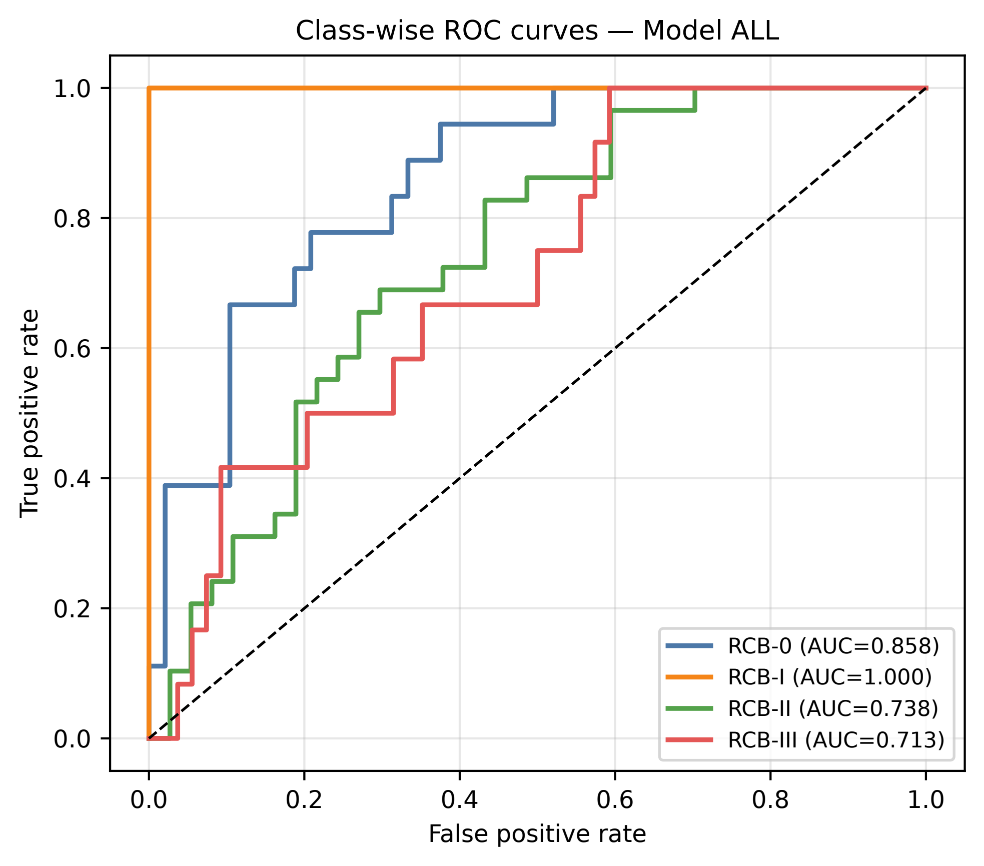

Class-wise ROC curves — Model ALL
True positive rate
False positive rate
0.0
0.2
0.4
0.6
0.8
1.0
RCB-0 (AUC=0.858)
RCB-I (AUC=1.000)
RCB-II (AUC=0.738)
RCB-III (AUC=0.713)

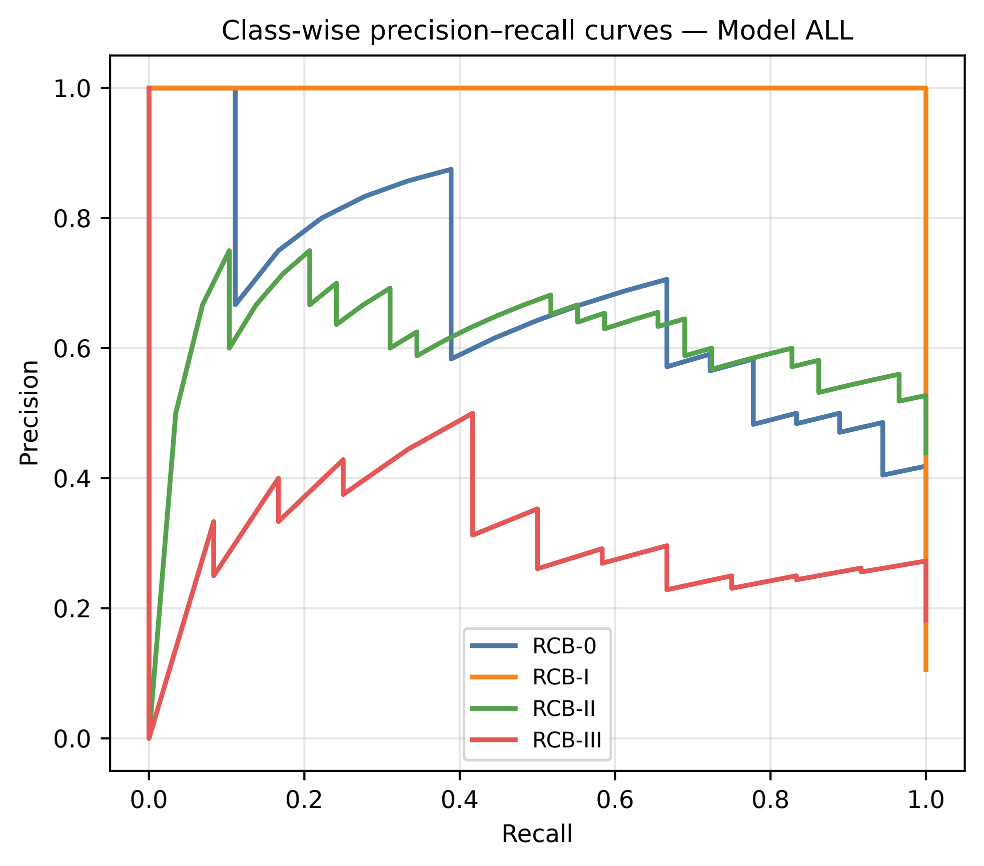

Class-wise precision–recall curves — Model ALL
Precision
Recall
1.0
0.8
0.6
0.4
0.2
0.0
0.0
0.2
0.4
0.6
0.8
1.0
RCB-0
RCB-I
RCB-II
RCB-III

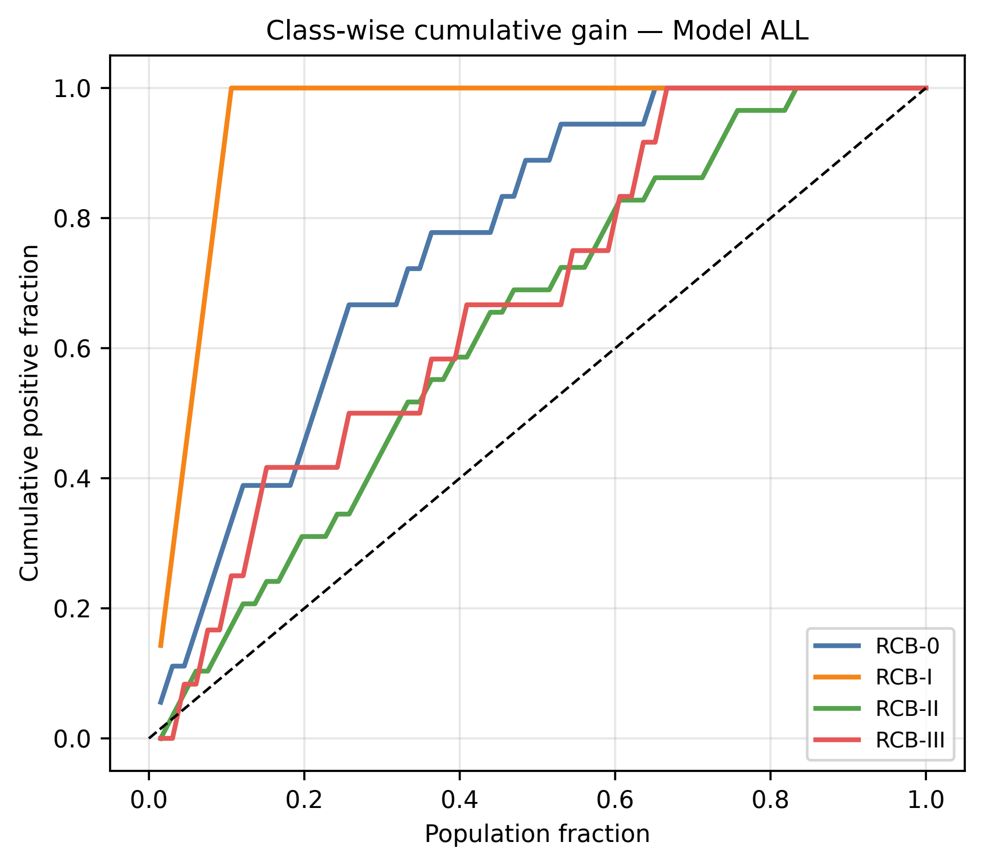

Class-wise cumulative gain — Model ALL
Cumulative positive fraction
Population fraction
0.0
0.2
0.4
0.6
0.8
1.0
RCB-0
RCB-I
RCB-II
RCB-III

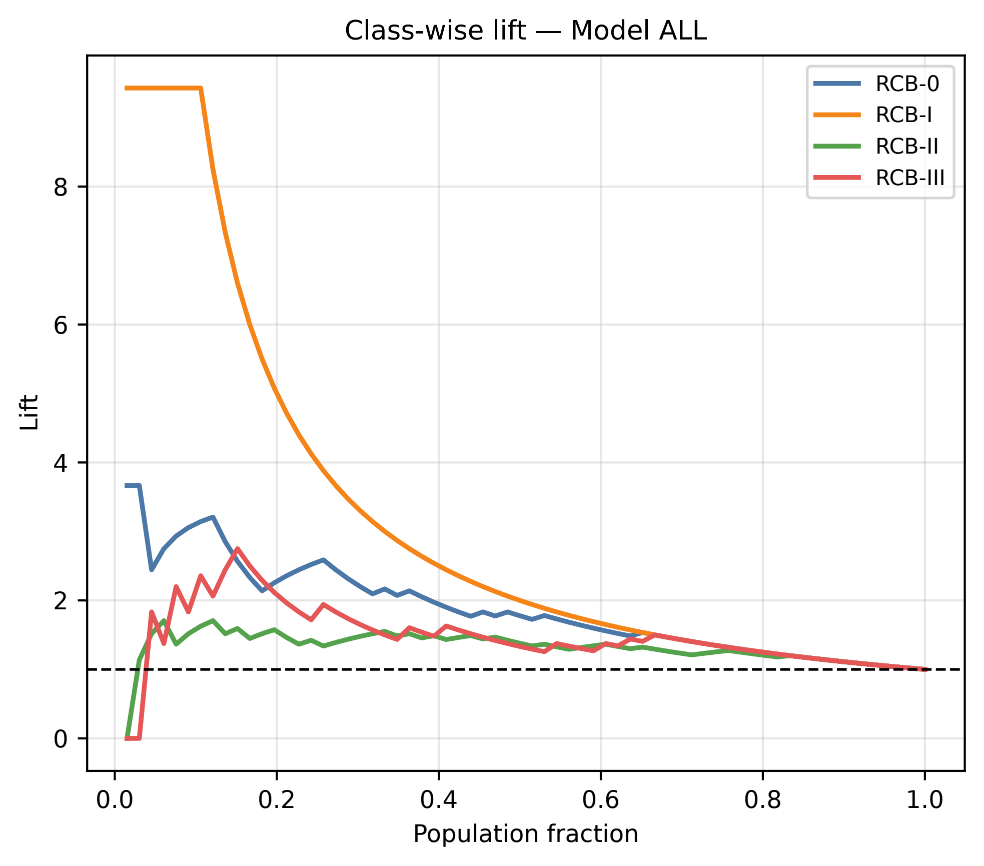
Class-wise lift — Model ALL
RCB-0
RCB-I
RCB-II
RCB-III
Lift
0
2
4
6
8
0.0
0.2
0.4
0.6
0.8
1.0
Population fraction

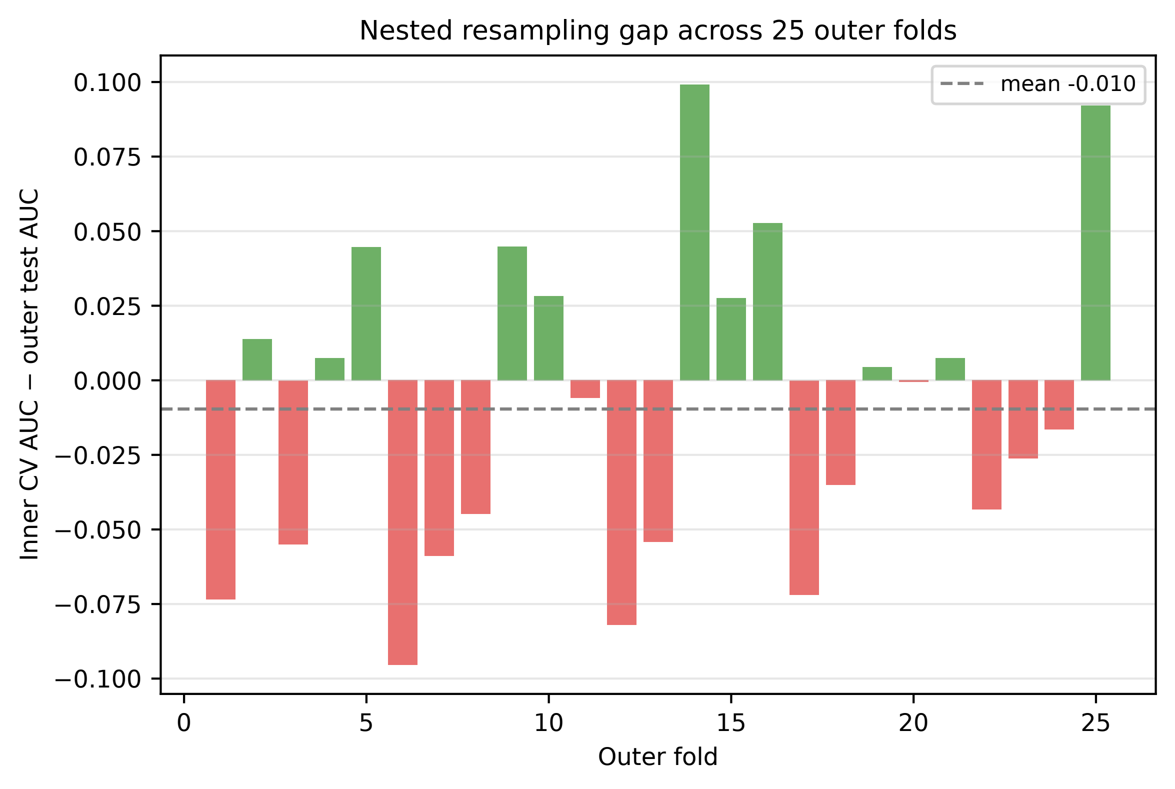

Nested resampling gap across 25 outer folds
mean -0.010
Inner CV AUC − outer test AUC
0.100
0.075
0.050
0.025
0.000
−0.025
−0.050
−0.075
−0.100
0
5
10
15
20
25
Outer fold

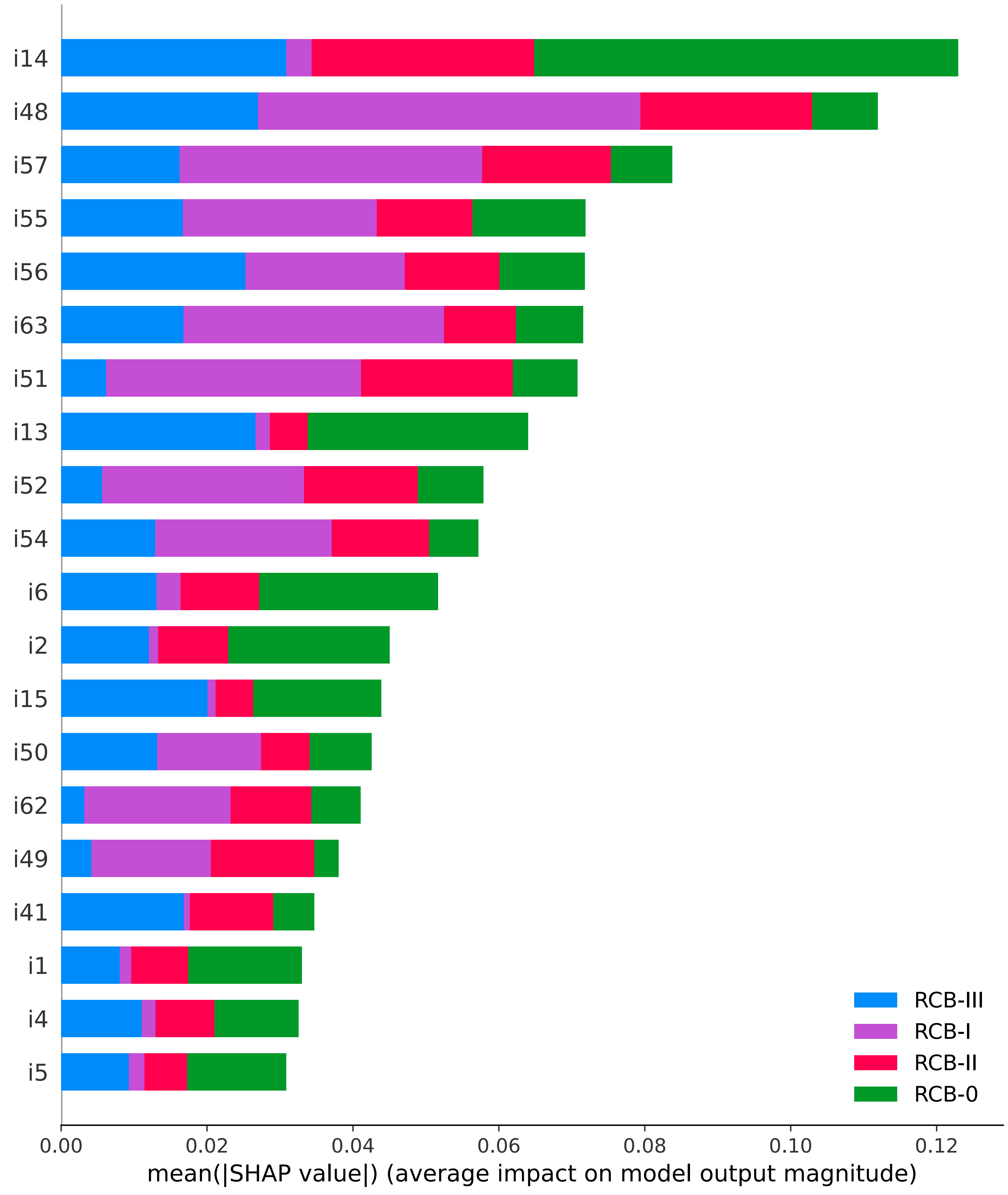

i14
i48
i57
i55
i56
i63
i51
i13
i52
i54
i6
i2
i15
i50
i62
i49
i41
i1
i4
i5
RCB-III
RCB-I
RCB-II
RCB-0
0.00
0.02
0.04
0.06
0.08
0.10
0.12
mean(|SHAP value|) (average impact on model output magnitude)

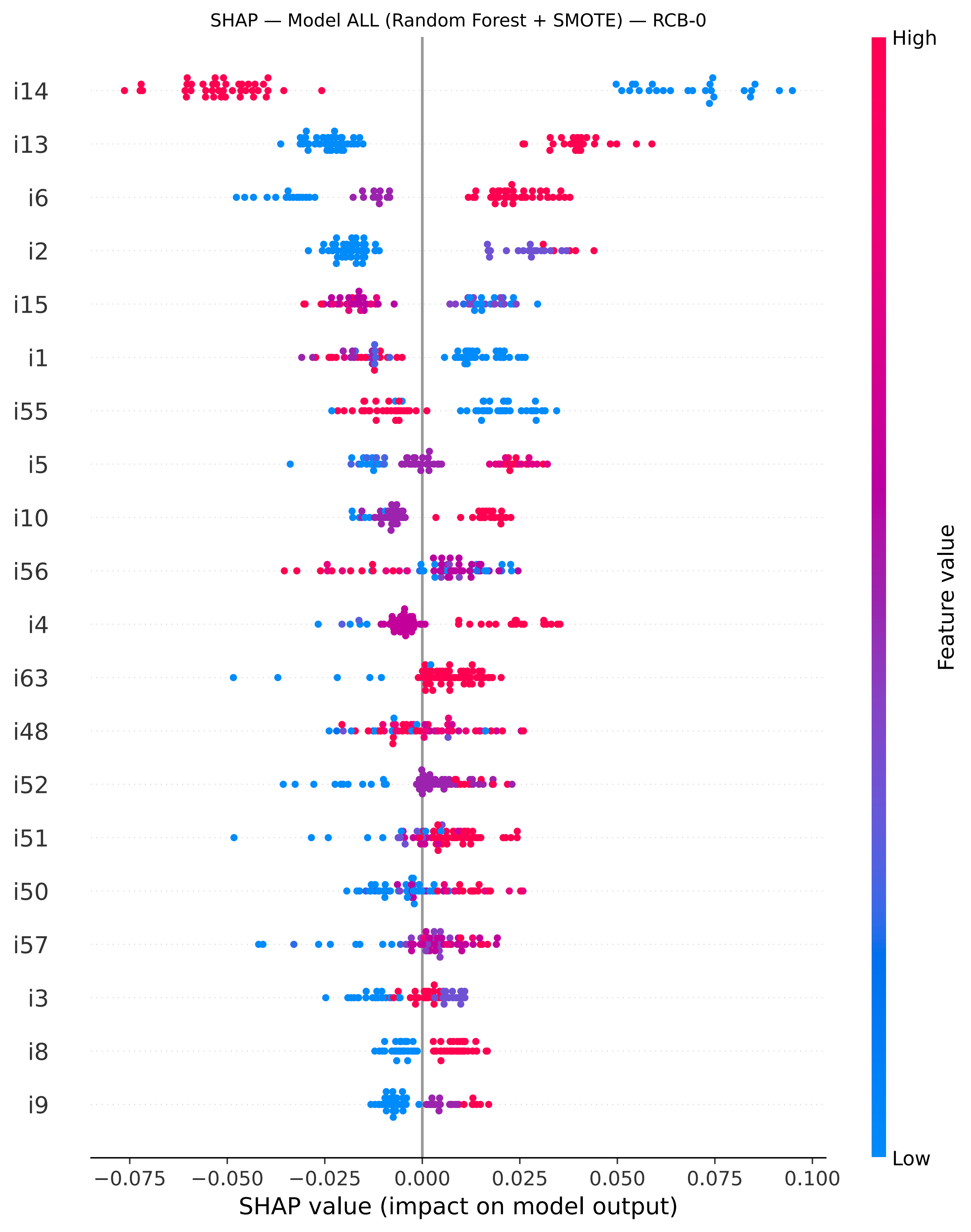

SHAP — Model ALL (Random Forest + SMOTE) — RCB-0
i14
i13
i6
i2
i15
i1
i55
i5
i10
i56
i4
i63
i48
i52
i51
i50
i57
i3
i8
i9
−0.075
−0.050
−0.025
0.000
0.025
0.050
0.075
0.100
SHAP value (impact on model output)
High
Low
Feature value

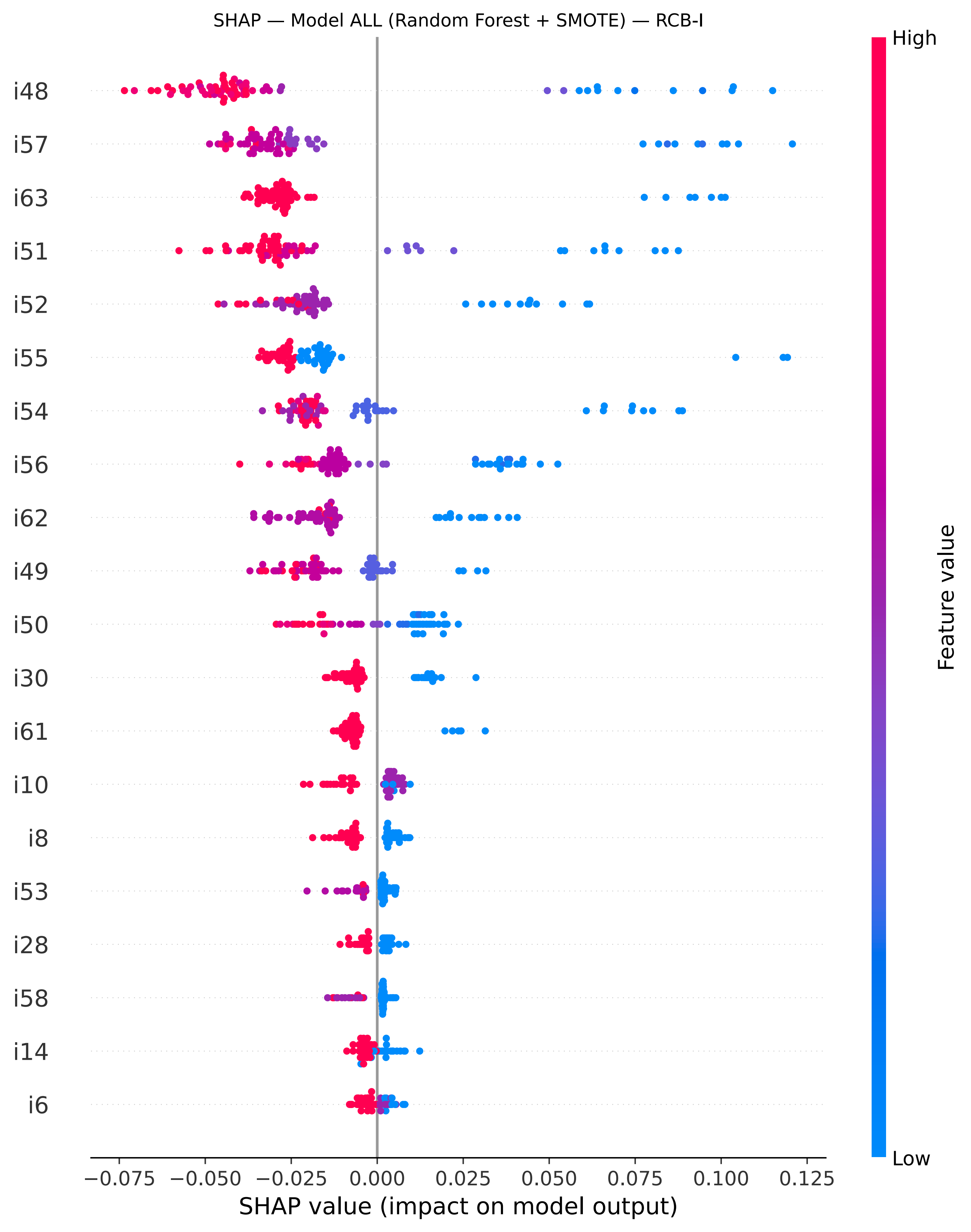

SHAP — Model ALL (Random Forest + SMOTE) — RCB-I
i48
i57
i63
i51
i52
i55
i54
i56
i62
i49
i50
i30
i61
i10
i8
i53
i28
i58
i14
i6
−0.075
−0.050
−0.025
0.000
0.025
0.050
0.075
0.100
0.125
SHAP value (impact on model output)
High
Low
Feature value

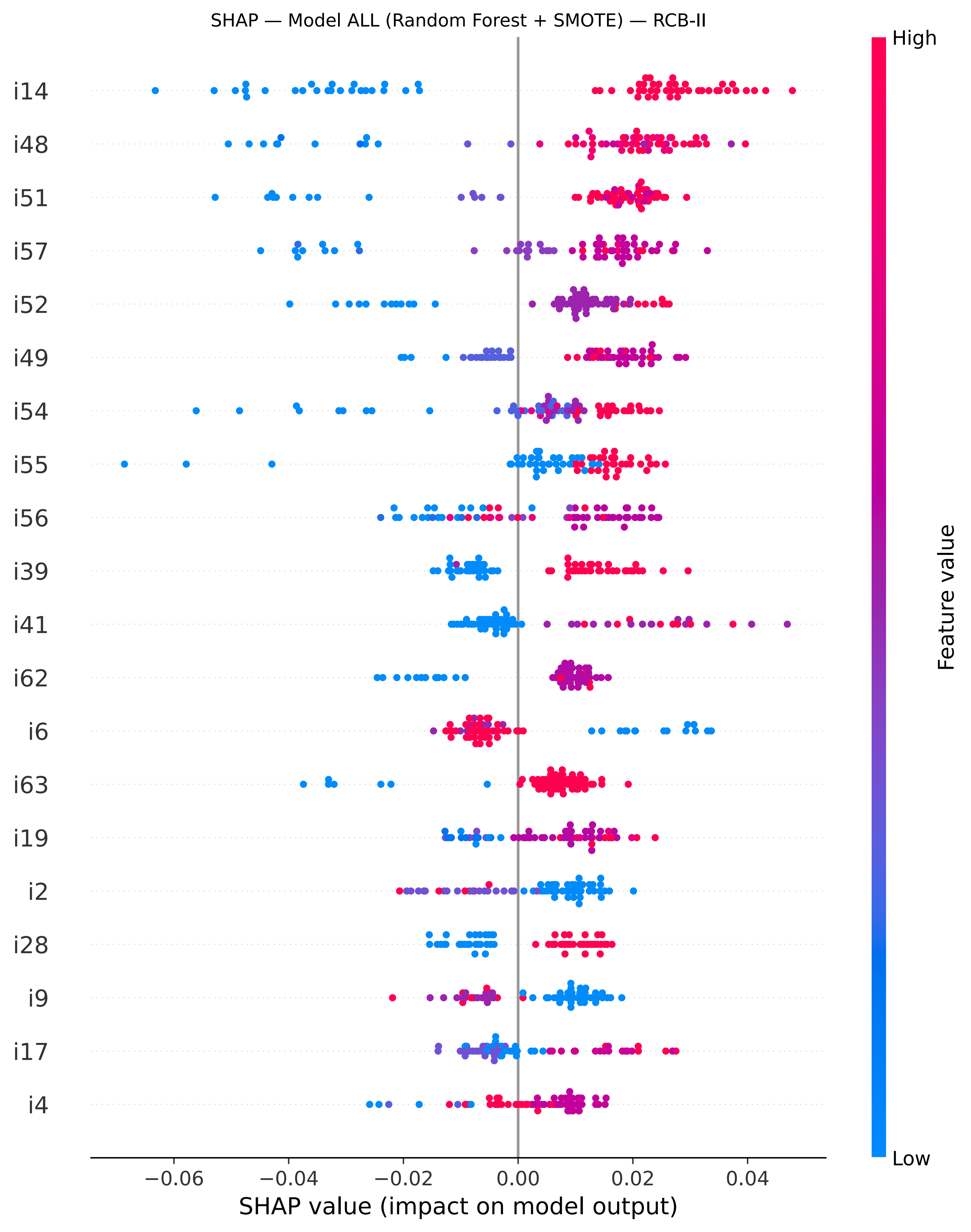

SHAP — Model ALL (Random Forest + SMOTE) — RCB-II
i14
i48
i51
i57
i52
i49
i54
i55
i56
i39
i41
i62
i6
i63
i19
i2
i28
i9
i17
i4
−0.06
−0.04
−0.02
0.00
0.02
0.04
SHAP value (impact on model output)
High
Low
Feature value

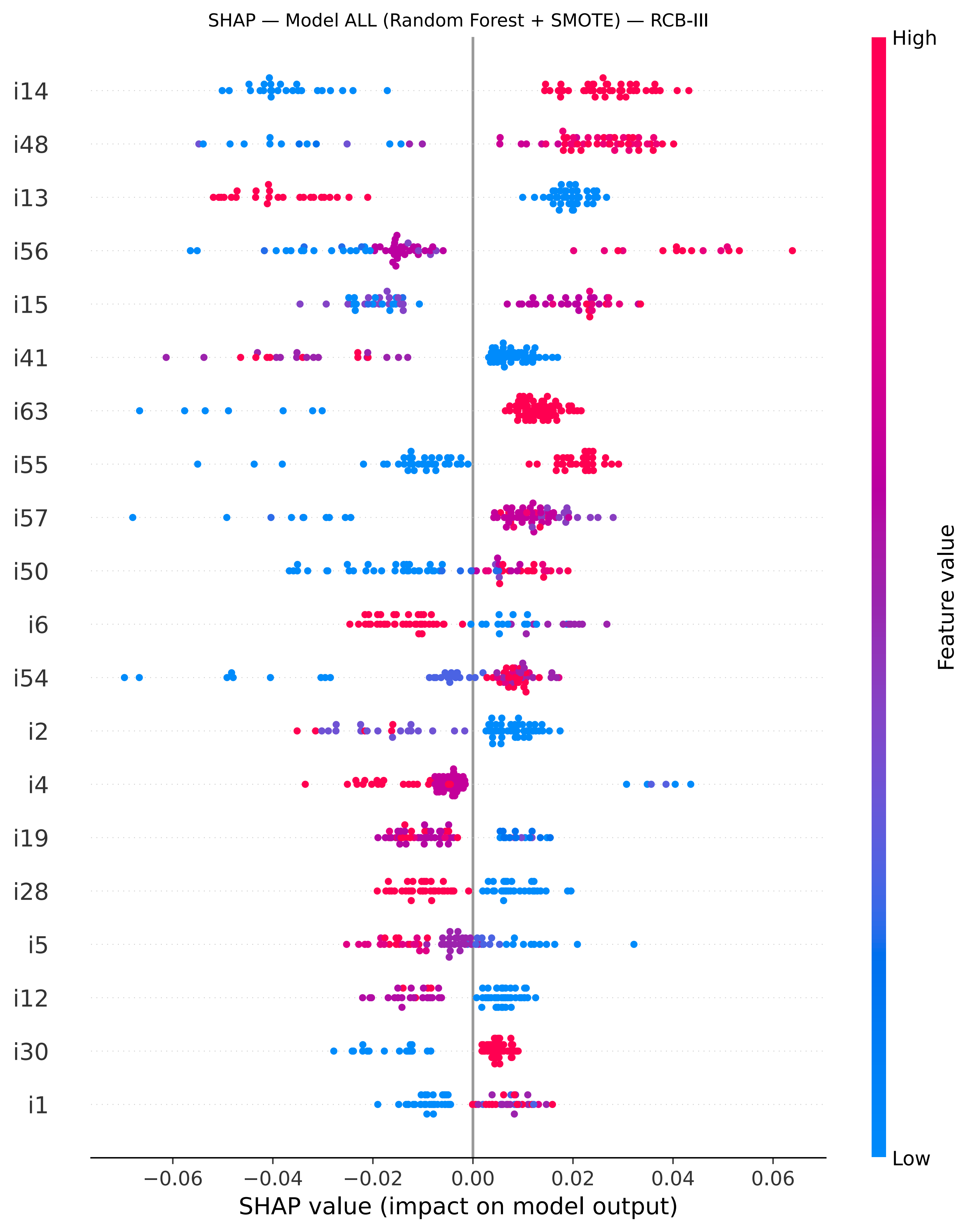
SHAP — Model ALL (Random Forest + SMOTE) — RCB-III
i14
i48
i13
i56
i15
i41
i63
i55
i57
i50
i6
i54
i2
i4
i19
i28
i5
i12
i30
i1
−0.06
−0.04
−0.02
0.00
0.02
0.04
0.06
SHAP value (impact on model output)
High
Low
Feature value

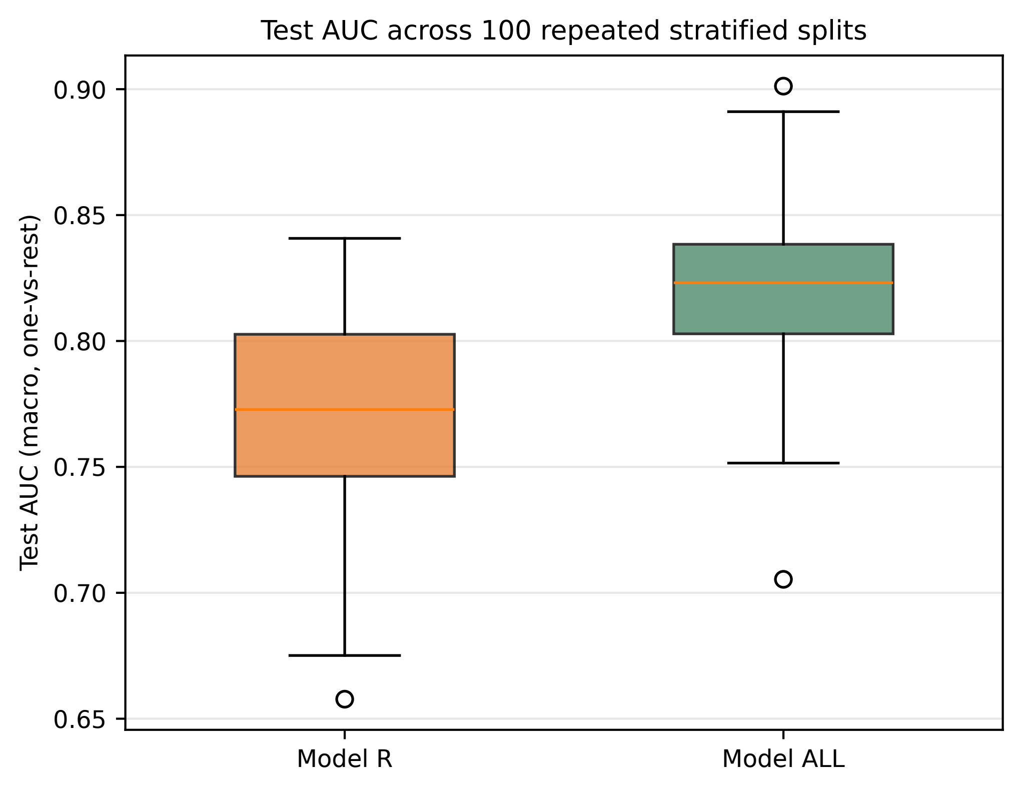
Test AUC across 100 repeated stratified splits
Test AUC (macro, one-vs-rest)
0.90
0.85
0.80
0.75
0.70
0.65
Model R
Model ALL

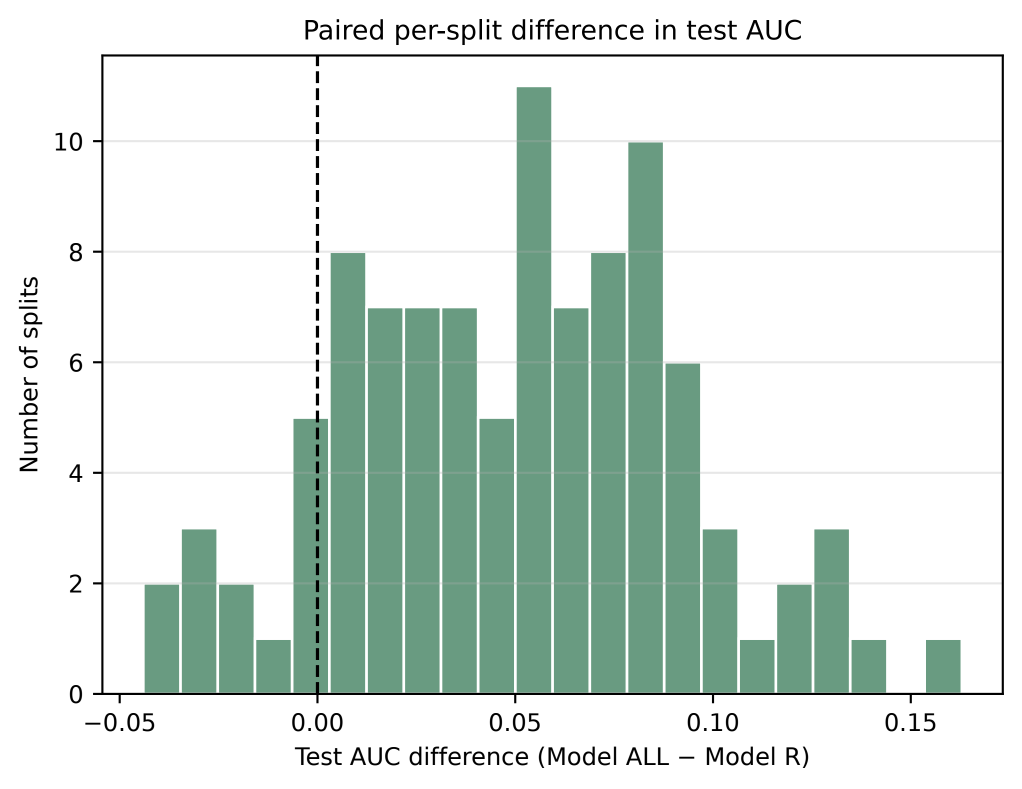
Paired per-split difference in test AUC
Number of splits
Test AUC difference (Model ALL − Model R)
0
2
4
6
8
10
−0.05
0.00
0.05
0.10
0.15

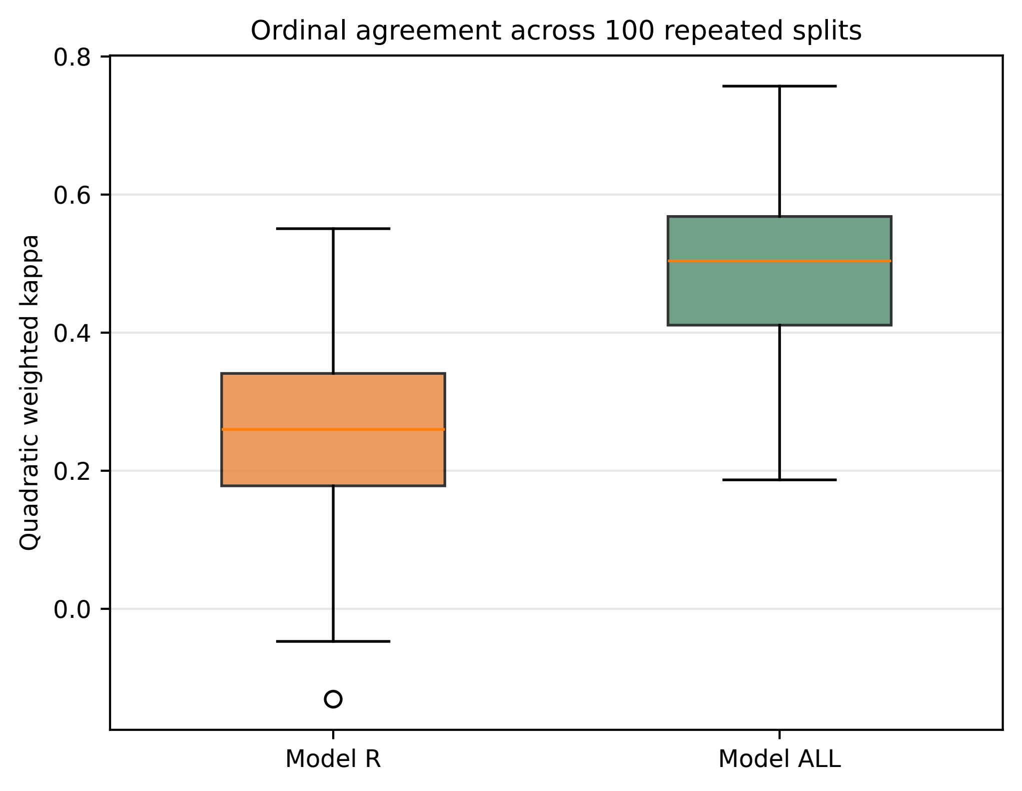

Ordinal agreement across 100 repeated splits
Quadratic weighted kappa
0.8
0.6
0.4
0.2
0.0
Model R
Model ALL

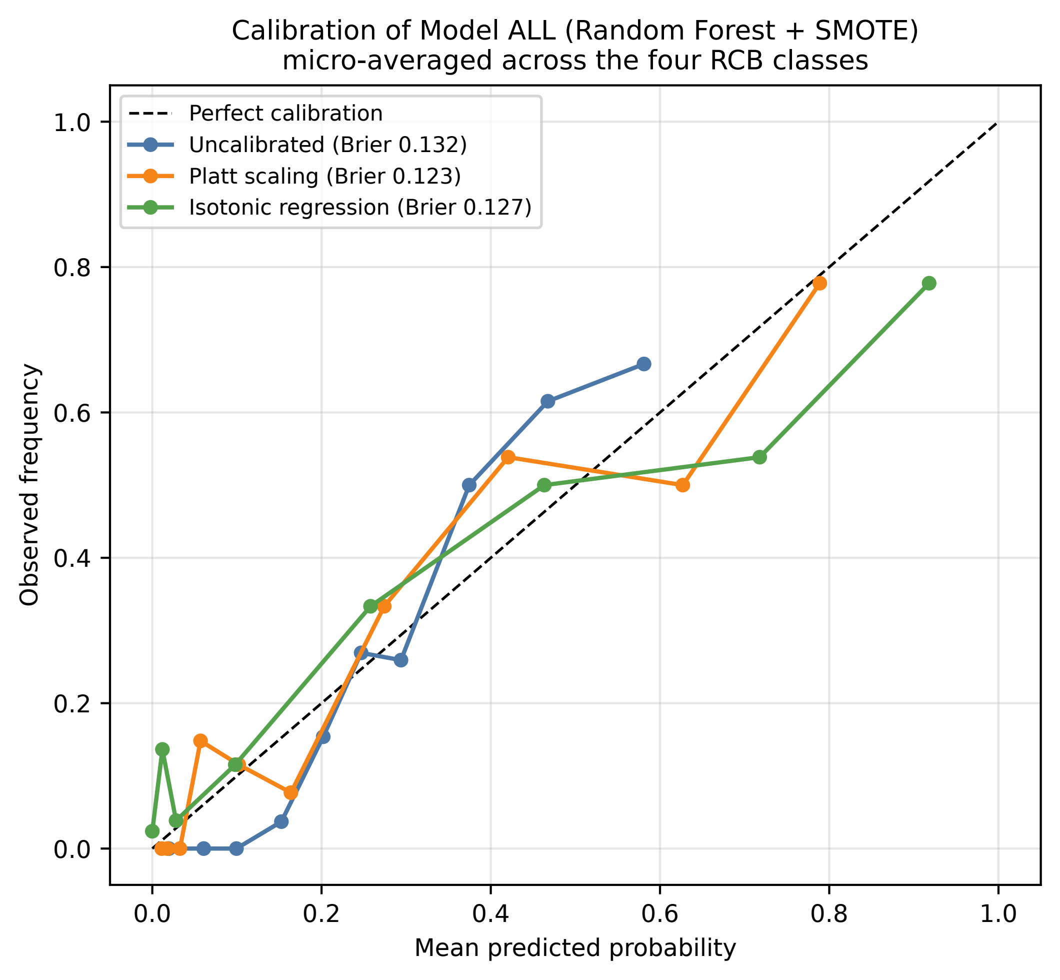

Calibration of Model ALL (Random Forest + SMOTE)
micro-averaged across the four RCB classes
Perfect calibration
Uncalibrated (Brier 0.132)
Platt scaling (Brier 0.123)
Isotonic regression (Brier 0.127)
Observed frequency
Mean predicted probability
0.0
0.2
0.4
0.6
0.8
1.0

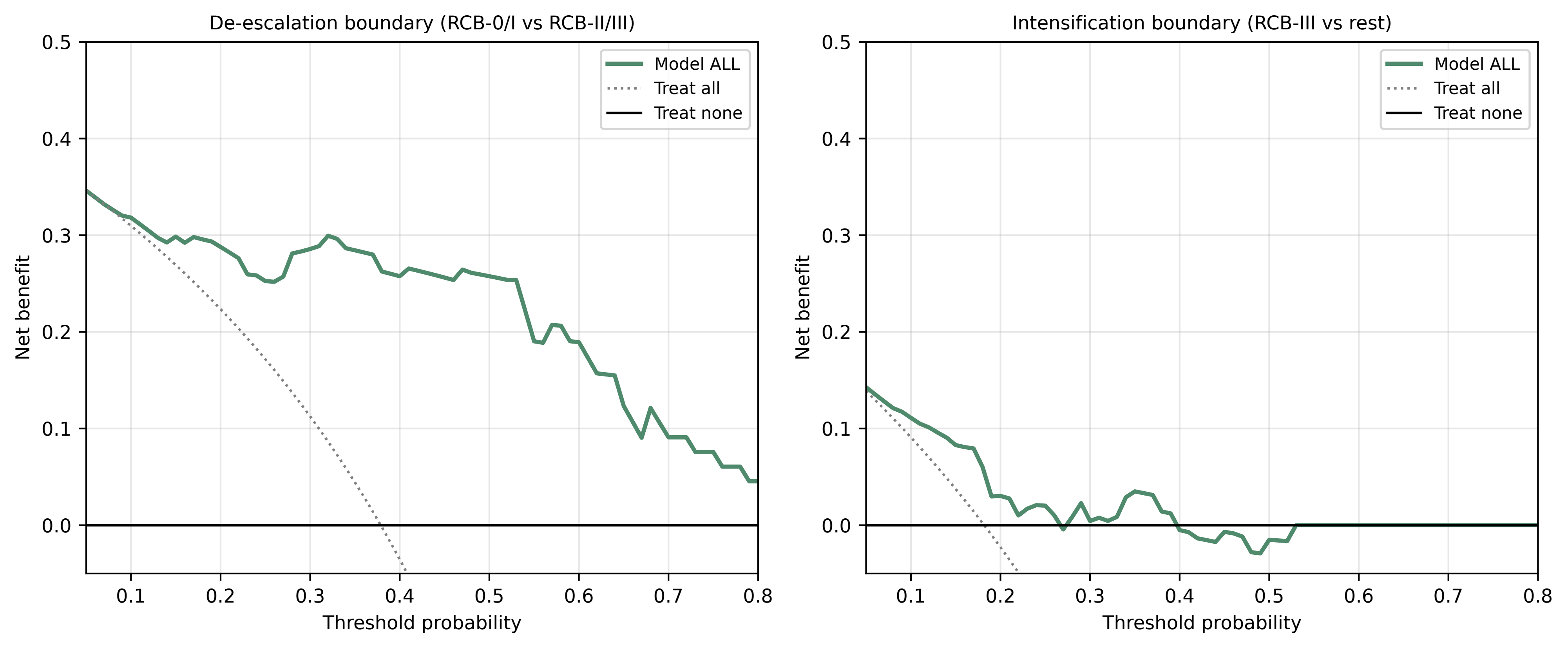

De-escalation boundary (RCB-0/I vs RCB-II/III)
Intensification boundary (RCB-III vs rest)
Model ALL
Treat all
Treat none
Net benefit
Threshold probability
0.5
0.4
0.3
0.2
0.1
0.0
0.1
0.2
0.3
0.4
0.5
0.6
0.7
0.8

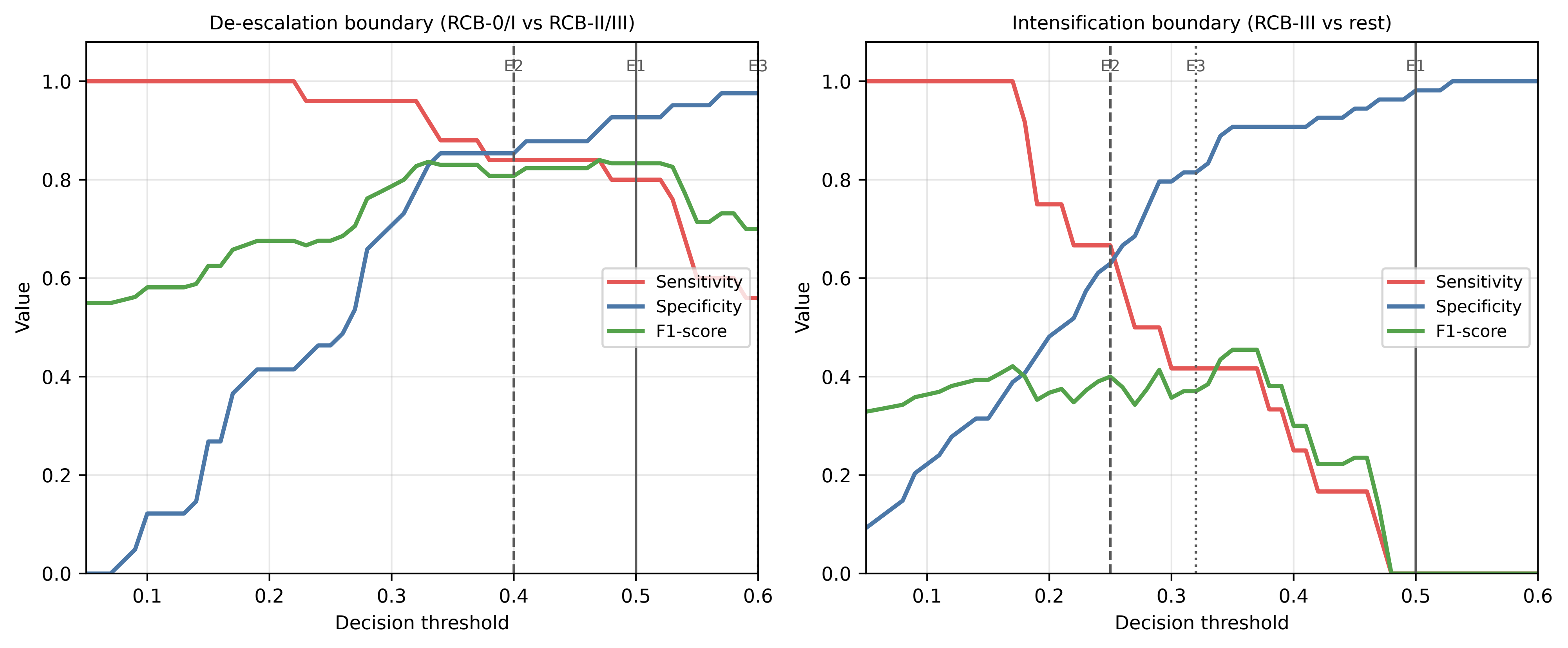
De-escalation boundary (RCB-0/I vs RCB-II/III)
E2
E1
E3
Sensitivity
Specificity
F1-score
Value
Decision threshold
Intensification boundary (RCB-III vs rest)
E2
E3
E1
Sensitivity
Specificity
F1-score
Value
Decision threshold
